# Oncogene Concatenated Enriched Amplicon Nanopore Sequencing for Rapid, Accurate, and Affordable Somatic Mutation Detection

**DOI:** 10.1101/2020.11.12.20230169

**Authors:** Deepak Thirunavukarasu, Lauren Y. Cheng, Ping Song, Sherry X. Chen, Mitesh J. Borad, Lawrence Kwong, Phillip James, Daniel J. Turner, David Yu Zhang

**Affiliations:** Department of Bioengineering, Rice University, Houston, TX; Department of Oncology, Mayo Clinic, Phoenix, AZ; Department of Translational Molecular Pathology, The University of Texas MD Anderson Cancer Center, Houston, TX; Oxford Nanopore Technologies, Oxford, UK; Department of Systems, Synthetic, and Physical Biology, Rice University, Houston, TX

## Abstract

Nanopore sequencing is more than 10-fold faster than sequencing-by-synthesis and provides reads that are roughly 100-fold longer. However, nanopore sequencing’s 7.5% intrinsic error rate renders it difficult to call somatic mutations with low variant allele frequencies (VAFs) without significant false positives. Here, we introduce the Oncogene Concatenated Enriched Amplicon Nanopore Sequencing (OCEANS) method, in which variants with low VAFs are selectively amplified and subsequently concatenated for nanopore sequencing. OCEANS allows accurate detection of somatic mutations with VAF limits of detection between 0.05% and ≤ 1%. We constructed 4 distinct multi-gene OCEANS panels targeting recurrent mutations in acute myeloid leukemia, melanoma, non-small-cell lung cancer, and hepatocellular carcinoma. Comparison experiments against Illumina NGS showed 99.79% to 99.99% area under the receiver-operator curve for these panels on clinical FFPE tumor samples. Furthermore, we identified a significant number of mutations below the standard NGS limit of detection in clinical tissue samples using each OCEANS panel. Comparison against digital PCR on 10 of putative mutations at ≤1% VAF showed 9 concordant positive calls with VAFs between 0.02% and 0.66%. By overcoming the primary challenge of nanopore sequencing on detecting low VAF single nucleotide variant mutations, OCEANS is poised to enable same-day clinical sequencing panels.

---

High-throughput DNA sequencing is becoming a standard part of oncology care, with many laboratory-developed tests informing patient prognosis [1], therapy selection[2, 3], and minimal residual disease [4]. DNA sequencing is furthermore being explored as a method for enabling early screening of cancers in asymptomatic populations [5, 6]. The dominant platforms used for clinical high-throughput sequencing today are based on sequencing-by-synthesis (NGS, e.g. the Illumina and Ion Torrent platforms).

Although NGS has many advantages including high throughput, high sensitivity, and high reproducibility/reliability, NGS also has three notable limitations: First, NGS read lengths are limited to roughly 300 nt, rendering it less suitable/sensitive to larger scale DNA alterations such as copy number variations and chromosomal translocations. Second, NGS is time-consuming, with multiple days needed for sequencing-by-synthesis chemistry, in addition to time- and labor-intensive library preparation and bioinformatic interpretation. Third, NGS requires a high capital investment on the order of $1,000,000 for a high-throughput instrument (e.g. Novaseq); more affordable NGS platforms such as the MiniSeq result in roughly 10-fold higher per-read sequencing costs.

Nanopore sequencing (NS) overcomes all three of the above limitations of NGS, with average read lengths of over 10,000 nt, sequencing times as short as 15 minutes, and Oxford Nanopore MinION instrument having list price of approximately $1,000 and size of a USB drive. On the other hand, NS is hamstrung by a different problem: high intrinsic single-pass sequencing error rates of about 7.5% [7], com-pared to less than 1% for NGS. Although there has been intense recent research effort in reducing NS error rates on both the hardware [8] and software [9] sides, calling somatic single-base mutations at variant allele frequencies (VAF) remaining challenging for NS due to its high single-molecule error rates [10–12].

A large fraction, if not a majority, of actionable cancer DNA alterations are single-base mutations [13], and in tumor tissue samples there may be significant cancer heterogeneity and/or low tumor fraction. Consequently, reliable detection of single-base mutations at 5% VAF in formalin-fixed, paraffin-embedded (FFPE) tissues is currently considered a standard requirement for clinical NGS assays [14]. Cell-free DNA in peripheral blood plasma is an emerging biospecimen for noninvasive cancer monitoring [15] and has even more stringent requirements on VAF limit of detection (LoD), with typical commercial NGS kits and services claiming LoDs of between 0.1% and 0.5% VAF.

Here, we present a new method, Oncogene Concatenated Enriched Amplicon Nanopore Sequencing (OCEANS), that enables NS to achieve ≤ 1% VAF LoD on single-base mutations in FFPE samples (Fig. 1). The OCEANS method integrates the blocker displacement amplification (BDA) allele enrichment method [16, 17] with a stochastic amplicon ligation (SAL), improving the VAF LoD by roughly 100-fold and throughput by roughly 10-fold over standard NS. The entire OCEANS method takes less than 10 hours from DNA to called variants, and the average sequencing cost per FFPE sample is roughly $3.75 for a 7-gene, 15-amplicon panel on the Oxford Nanopore MinION flow cells.

**FIG. 1:**
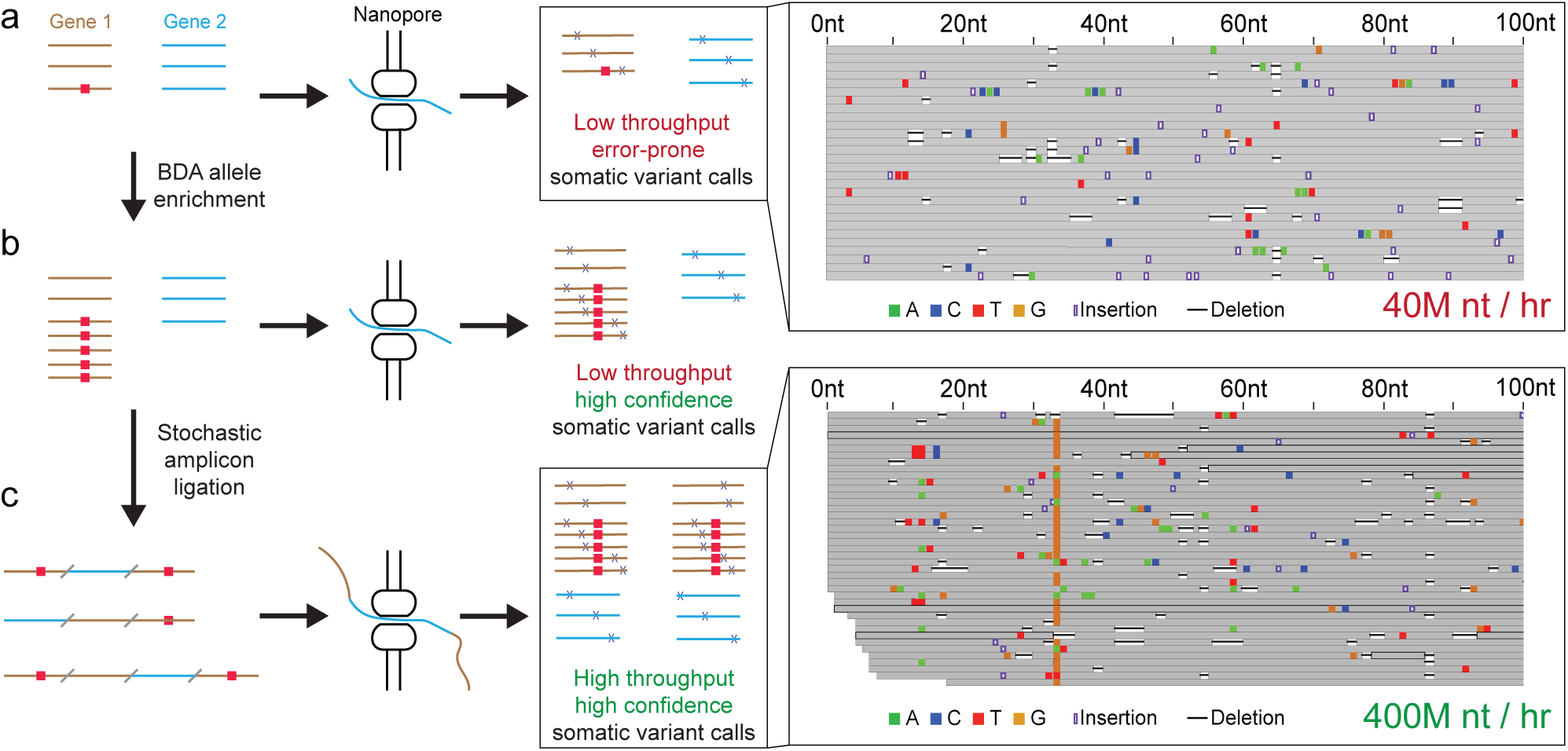
Overview of Oncogene Concatenated Enriched Amplicon Nanopore Sequencing (OCEANS) approach. **(a)** Short DNA genes or loci of interest potentially bearing somatic mutations (red rectangle) are inefficiently and inaccurately sequenced using nanopore sequencing (NS). The high error rate of NS renders it difficult to confidently call somatic mutations with less than 50% variant allele frequency (VAF), and the short lengths of DNA from oncology biospecimen (e.g. FFPE, cfDNA) reduce the throughput of NS. **(b)** We first use the blocker displacement amplification (BDA) technology to selectively amplify DNA sequence variants, so that somatic mutations with low sample VAF (≤ 5%) are represented in high VAF in the prepared DNA library. **(c)** Subsequently, we enzymatically concatenate the amplicons to increase the effective throughput of NS. Compared to standard NS, OCEANS exhibits roughly 100-fold better mutation VAF limit of detection and roughly 10-fold higher throughput.

## Results

### Stochastic Amplicon Ligation

DNA samples for oncology sequencing are typically extracted from FFPE tissues, and can have average lengths of less than 500 nt due to accumulated chemical damage [18]. This presents a unique challenge for nanopore sequencing (NS) because NS yields roughly the same number of reads in the first hour of sequencing regardless of the length of each read (Fig. 2a). This means that the throughput of NS in nucleotides for FFPE-derived DNA is only a small fraction of the flow cell’s achievable potential. To overcome this challenge, we developed the Stochastic Amplicon Ligation (SAL) method to enzymatically concatenate many short DNA molecules together to increase the effective throughput of NS.

**FIG. 2:**
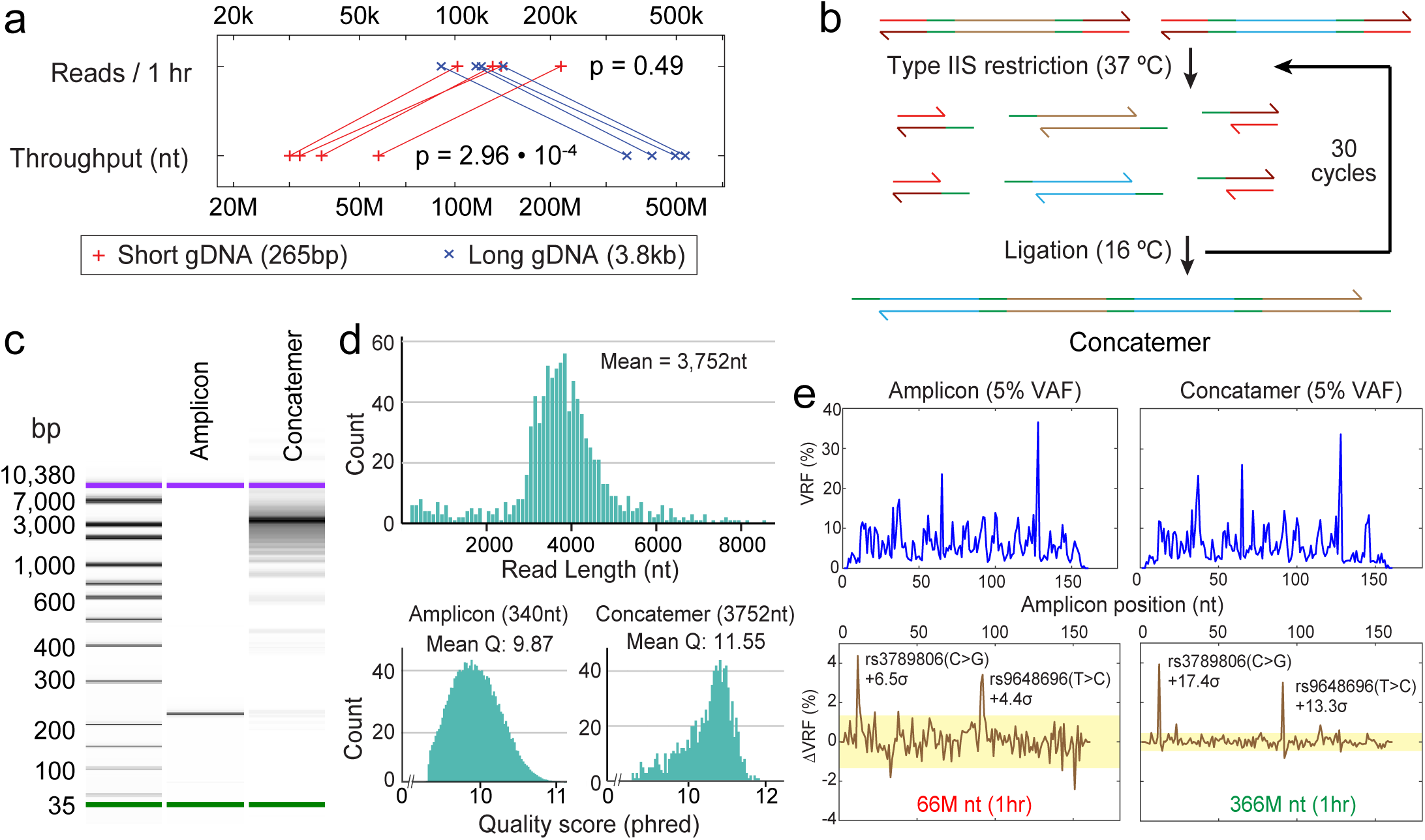
Concatenation of short amplicons by Stochastic Amplicon Ligation (SAL). **(a)** The number of NS reads per hour is observed to be roughly invariant with the length of the DNA molecules in the sequencing library. Here, we performed NS on the NA18562 human genomic DNA (gDNA) physically sheared (Covaris g-tube) to an average of 265 base pairs (bp) or 3,800 bp (3.8 kb). The number of reads in 1 hour of sequencing on an Oxford Nanopore MinION flow cell was observed to be similar for the two libraries (*p* = 0.49), indicating that the effective throughput of the longer gDNA library was roughly 10-fold higher. **(b)** Concatenation of DNA amplicons through SAL. Amplicons are adapted so that their 5^*′*^ and 3^*′*^ ends possess Type IIS restriction enzyme sites. Through a series of restriction and ligation reactions, >10 amplicons are assembled into concatemers. Compared to a naive method based on blunt end ligation, SAL enriches longer concatemers by maintaining low concentrations of amplicon monomers during each cycle of the reaction. Additionally, SAL significantly increases the on-target rate of the final NS libraries by excluding undesired dsDNA molecules from being incorporated into the concatemer. **(c)** Capillary electrophoresis analysis of a 220 nt amplicon and its SAL concatemer products. **(d)** NS read lengths of concatemers for a 7-plex SAL reaction from amplicons with a mean length of 340 nt. In addition to the increased throughput of NS for the concatemer due to the longer DNA lengths, we also observed a significant increase in sequencing quality for the concatemer vs. the original amplicon. **(e)** Increased NS throughput from SAL improves the limit of detection of somatic mutations when paired normal samples are available. The top traces show the variant read fraction (VRF) of NS reads at each location that corresponds to the highest frequency single-base changes (substitution, insertion deletion). The bottom traces show the relative excess of the top variant at each position for the 5% VAF samples compared to a 0% VAF sample; see Supplementary Section S2 for amplicon NS traces for 0% VAF samples. The two SNP alleles specific to NA18562 were more prominently called in the SAL concatemer NS results. The input DNA for each run was either 50 ng of a 95%:5% mixture of the NA18537 and NA18562 cell line human genomic DNA (gDNA), or 50 ng NA18537.

SAL is based on the Golden Gate assembly method used in synthetic biology to concatenate short oligos into synthetic genes [19]. In SAL (Fig. 2b), amplicons are appended with engineered adapter sequences that possess a Type IIS restriction enzyme recognition site. After Type IIS cleavage, a 4 nt sticky end is left on the 5’ ends of both strands of the amplicons; these sticky ends allow the amplicons to transiently bind to each other, which then enzymatically ligate to form concatemers. Multiple cycles of enzymatic restriction and ligation are performed to increase the lengths of the concatemers, and we perform a SPRI size selection afterwards to both enrich long concatemers and remove short recognition site oligos cleaved from the amplicons. The multiple temperature cycles between 37 ^*°*^C and 16 ^*°*^C improve the mean lengths of the concatemer assemblies by keeping the concentrations of activated monomer amplicons low. Simulations and prior literature [19] suggest that direct ligation of amplicons with 5^*′*^ sticky ends results in a much larger population of shorter concatemers.

SAL differs from traditional Golden Gate assembly in having universal sticky end sequences to allow stochastic incorporation of any amplicon with the appropriate adapters. This, in principle, allows unlimited growth of concatemers to longer lengths given sufficient monomer concentration and enough temperature cycles, and reduces the possibility of unintended reactions due to nonspecific binding between non-cognate sticky ends. Experimentally, capillary electrophoresis indicated that SAL concatenated a mean of roughly 12 to 15 monomers per concatemer (Fig. 2c). In addition to improving the throughput of NS, we also found that the SAL reduced the quality/error rate of NS (Fig. 2d). The NS results of a 340 nt amplicon had a mean phred quality score of 9.87, corresponding to an error rate of 10.3%. The concatemer, in contrast, had a mean phred score of 11.55, corresponding to an error rate of 7.0%.

The improved throughput and accuracy of NS of SAL concatemers allow calling somatic single-base mutations at 5% VAF when matched normal sample are available (Fig. 2e). We first applied NS to amplicons from a 95%:5% mixture of the NA18537 and NA18562 human cell line genomic DNA. NA18537 and NA18562 were homozygous for different alleles at the rs3789806 and rs9648696 single nucleotide polymorphism (SNP) loci, so the mixture was 5% VAF in the NA18537 SNP alleles. The high error rates of NS resulted in a large number of loci on the amplicon with high variant read fre-quencies, which would all be false positive variant calls if we attempted to call somatic mutations at 5% VAF. When we subtracted the variant read fraction (VRF) from a 100% NA18537 sample, then the 5% VAF somatic mutations became more visible in the ΔVRF. In direct amplicon NS, the two 5% VAF SNP alleles were detected in the ΔVRF figure with +6.5*σ* and +4.4*σ*, respectively. The confidence of calling these 5% VAF variants were increased to +17.4*σ* and +13.3*σ* for SAL concatemers. For SAL concatemers, the long NS reads were bioinformatically deconcatenated using a custom python code (Supplementary Section S3).

Rolling circle amplification (RCA) is an alternative method for generating long DNA from shorter DNA molecules, which has been used in the context of NS for improving accuracy [20]. RCA has several significant limitations compared to SAL, most notably that RCA requires an initial circularization of DNA which is known to have low efficiency, reducing the clinical sensitivity due to low conversion yield of biological DNA molecules in sequencing library. Additionally, RCA produces concatemers in which the segment sequences all reflect the sequence of the original molecule, rather than a uniform sampling of all DNA molecules on the loci of interest. Finally, RCA generates single-stranded DNA products rather than double-stranded DNA products, which should be converted into double-stranded DNA for efficient NS.

### Integrating BDA Allele Enrichment

Frequently, matched normal FFPE tissue samples will not be available, so using SAL alone for NS detection of low VAF so-matic mutations is unlikely to be impactful clinically. The OCEANS method employs blocker displacement amplification (BDA) [16, 17] to allow more robust detection of low VAF somatic mutations without requiring a matched normal sample. In brief, BDA includes a wildtype-binding blocker oligonucleotide that competes with a PCR primer in hybridizing to DNA templates of interest. The blocker binds more strongly than the primer to wildtype DNA sequences, preventing efficient PCR amplification. On DNA templates with sequence variants, the primer outcompetes the blocker and PCR proceeds as usual. Through the course of many PCR cycles, the VAF of sequence variants (including single nucleotide mutations) can be enriched by over 1000-fold.

In OCEANS, the DNA biospecimen is first mixed with multiple primers and blockers to undergo variant-selective PCR amplification (Fig. 3a). The amplicons will over-represent sequence variants in genetic loci of interest, though some amplicons with wildtype sequences will still exist. The amplicons are subsequently appended with SAL adapters and concatenated into concatemers, and size-selected to remove short assemblies, primers, etc. The concatemers are then ligated to the standard Oxford Nanopore sequencing adapters with attached motor proteins, and loaded into the NS flow cell. The entire workflow takes about 10 hours, including post-sequencing bioinformatics. On the same SNP alleles as in Fig. 2, the OCEANS results showed VRFs that were dramatically higher than the sample variant VAFs (Fig. 3b), with 0.1% VAF SNP allele enriched to over 70% VRF. Thus, OCEANS allows robust variant calls of somatic mutations without the need for a matched normal DNA sample, which was not possible previously on the nanopore sequencing platform.

**FIG. 3:**
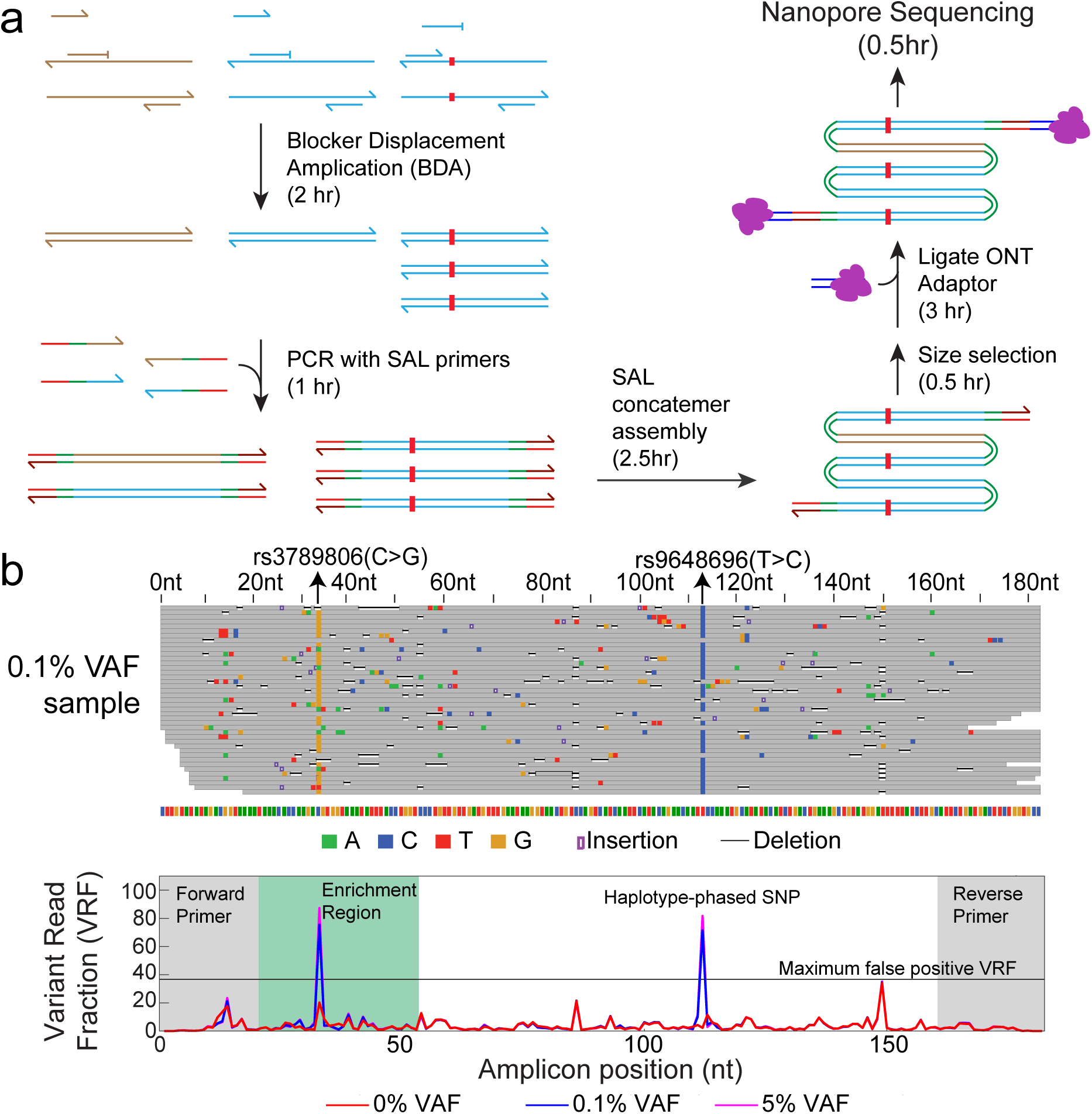
Method and experimental results for the full OCEANS method. **(a)** Potential variants in multiple genetic loci of interest are first enriched using multiplex blocker displacement amplification (BDA) [16, 17]. The amplicons are subsequently appended with SAL adapters using PCR, and assembled into concatemers. After size selection to remove short concatemers and excess primers, the concatemers are ligated to the Oxford Nanopore adapter bearing a motor protein, and sequenced using the MinION platform. **(b)** OCEANS enables confident variant calls of single-base variants at 0.1% VAF without a matched normal sample. The top diagram shows a randomly selected subset of NS reads, and the bottom diagram shows the variant read frequency (VRF), the fraction of NS reads at each locus that corresponds to the most frequent single-base variant. The forward and reverse primer regions are shaded in gray, and the BDA enrichment region is shaded in green.

We next constructed two multiplexed OCEANS panels: a 7-amplicon panel covering recurrent mutations observed in acute myeloid leukemia (AML) and a 15-amplicon panel covering recurrent mutations observed in melanoma. The AML panel covers roughly 254 mutations in the COSMIC database across 7 genes (Fig. 4a), and the melanoma panel covers roughly 370 mutations across 8 genes (Fig. 4d) We first characterized the limit of detection for mutations covered by these OCEANS panels using synthetic spike-in reference samples, with VAFs ranging from 0.05% and 1%.

**FIG. 4:**
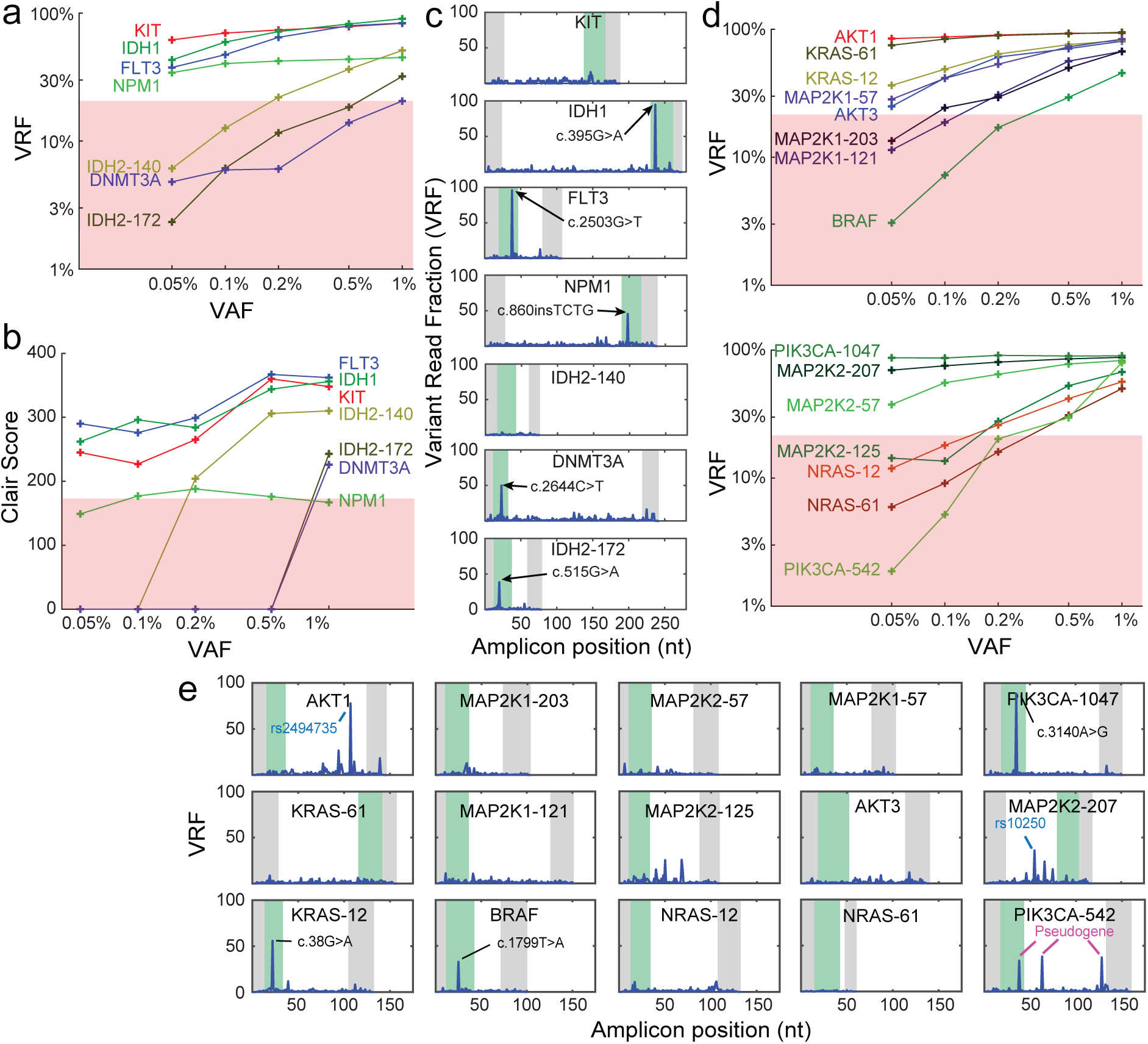
Bioinformatics and limit of detection of somatic mutation calls from multi-gene OCEANS panels. **(a)** Summary of variant read frequencies (VRF) observed for a 7-plex OCEANS panel covering recurrent mutations in the KIT, IDH1, FLT3, NPM1, IDH2, and DNMT3A genes, which are present in high frequency in acute myeloid leukemia (AML). The input samples used here were internal reference samples constructed by spiking known quantities of synthetic DNA bearing a mutation of interest into NA18562 gDNA; see Supplementary Section S4 for summary of NS results on these experiments. Based on calibration experiments, we tentatively propose a 20% VRF cutoff for calling a variant. The limit of detection for different mutations varied between ≤0.05% VAF (for KIT, IDH1, FLT3, and NPM1) and 1% VAF (for DNMT3A). **(b)** Qualitative *de novo* variant calls using the Clair [21] bioinformatics pipeline, on the same data as in panel (a). Based on Oxford Nanopore internal calibration, Clair scores ≥180 are reliable. Although the Clair variant call differed from the VRF-based variant call for the NPM1 mutation, all other mutations were detected with similar VAF limits of detection. To be conservative and reduce false positive variant calls, we typically make a somatic variant call only when both VRF ≥20% and Clair ≥ 180. **(c)** Results of the AML 7-plex OCEANS panel on 50ng of a third-party reference DNA sample (Horizon HD829). All mutations in the reference sample covered by our panel were at 5% VAF, and were detected. **(d)** Summary of VRF observed for different internal reference samples using a 15-plex OCEANS panel covering 7 genes frequently mutated in melanoma (AKT1, AKT3, KRAS, MAP2K1, MAP2K2, NRAS, and PIK3CA). For this panel, the limits of detection varied between ≤0.05% VAF and 0.5% VAF. **(e)** Results of the melanoma 15-plex OCEANS panel on 50ng of a 99%:1% mixture of NA18562 gDNA and a third-party reference DNA sample (Horizon HD238). The HD238 sample is nominally a reference sample that is 50% VAF in BRAF-V600E (c. 1799T>A), so the input sample should be positive only for BRAF-V600E at 0.5% VAF. Upon our discovery of the KRAS-c.38G>A and PIK3CA-c.3140A>G mutations, we checked with the manufacturer and confirmed that these mutations are also present in the reference sample at low VAFs.

Variant calls were made using two different approaches: (1) based on the variant read frequency exceeding a threshold of 20%, and (2) based on a Clair [21] score of above 180. We found that both approaches were imperfect: considering VRF alone ignores the fact that NS has different error rates for certain sequences, e.g. homopolymers. Clair scores, on the other hand, are not monotonic with VAF and have been observed to be less accurate for indel calls [10, 25]. To ensure minimal false positives in variant calls, we require that a variant must be independently called by both methods in order to be reported. On our internal reference samples (Fig.4abd), we observed VAF limits of detection between 0.05% and 1%. We next applied our OCEANS panels to third-party reference samples from Horizon Discovery, with mutation VAFs at 5% (for AML) and 0.5% (for melanoma). The expected mutations were all called.

Interestingly, we also made a number of unexpected variant calls in the melanoma OCEANS panel (Fig. 4e). The variant in the MAP2K2-207 amplicon was confirmed to be a non-pathogenic SNP (rs10250). The variants in the PIK3CA-542 amplicon were found to be aligned to the PIK3CA pseudogene (LOC100422375) [22], which were also preferentially enriched by BDA. The “variants” associated with the PIK3CA pseudogene were bioinformatically excluded from variant calls in subsequent experiments. Finally, we made confident variant calls for PIK3CA c.3140A>G and KRAS c.38G>A. We contacted Horizon Discovery customer support regarding these putative mutations, and the latter confirmed that these mutations are also present at low VAFs in the HD238 sample.

### Validating OCEANS on Clinical Tissue Samples

We next applied the melanoma OCEANS panel to clinical melanoma tissue samples, including both fresh/frozen (FF) and FFPE tissue (Fig. 5ab). As in the calibration experiments, we called somatic mutations only when the VRF was observed to be greater than 20%, and the Clair score was above 180. In total, DNA from 7 FF and 18 FFPE tissue samples were sequenced using both OCEANS and NGS. The melanoma OCEANS panels cover a total of 384 loci, corresponding to a total of 9600 total loci analyzed across the 25 samples.

**FIG. 5:**
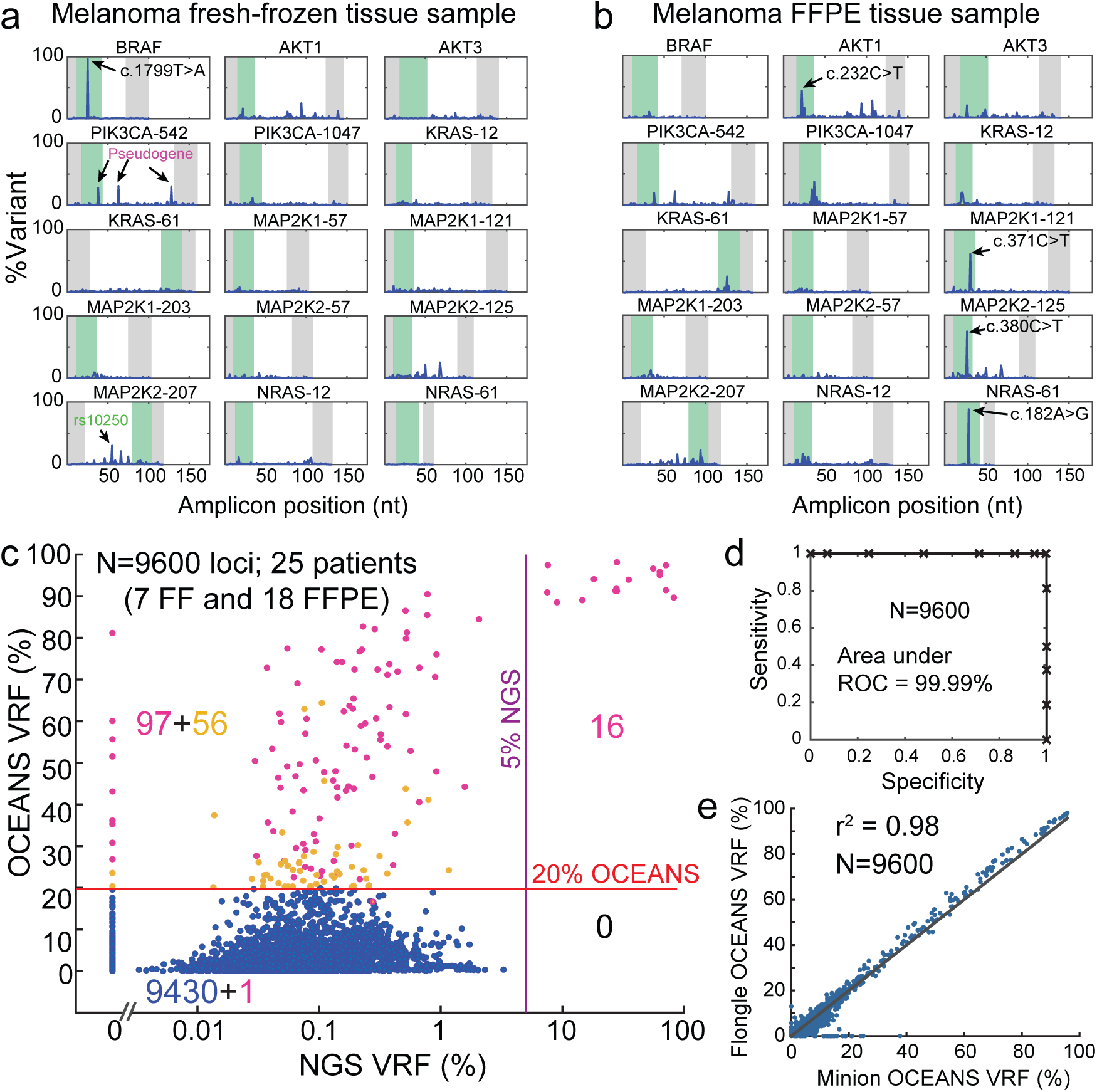
Validation of the 15-plex OCEANS melanoma panel on melanoma tissue samples. **(a)** Example results from a fresh-frozen melanoma tissue sample. The BRAF-V600E mutation was the only mutation called for this sample. **(b)** Example results from a formalin-fixed, paraffin-embedded (FFPE) tissue sample. 5 mutations were called in the AKT1, MAP2K1, MAP2K2, and NRAS genes. Other variants with ≥20% VRF that were not called by Clair are not labeled in the figure. All 5 of these mutations had confirmatory reads on a parallel NGS experiment, but only the NRAS c. 182A>G mutant had a NGS VRF of above 5%; see Supplementary Section S5 for details. **(c)** Summary of sequencing results for 25 clinical melanoma tissue samples (7 fresh/frozen, 18 FFPE). Input DNA quantities ranged from 10 ng and 50 ng (Supplementary Section S5). The X-axis shows the VRF based on a standard NGS analysis, and the Y-axis shows the OCEANS VRF. The horizontal line shows the 20% VRF cutoff for OCEANS variant calls, and the vertical line shows the 5% VRF cutoff for NGS variant calls. The numbers in quadrants display the number of loci in each group; 97 of the 153 putative variants in the top-left quadrant also had a Clair score of above 180 (purple dots). Relative to the NGS results, the OCEANS panel had a sensitivity of 100% and a specificity of 99.0%. Importantly, we believe that many of the 97 NGS-negative and OCEANS-positive results were true mutations, and our ddPCR confirmation experiments support this hypothesis (Table 1). We did not observe any significant difference for fresh/frozen samples vs. FFPE samples (Supplementary Section S5). **(d)** Receiver operator characteristic (ROC) curve for data in panel (c), based on changing the VRF cutoff for OCEANS. The area under the curve (AUC) is 99.99%. **(e)** High concordance of OCEANS results using Oxford Nanopore MinION vs. Flongle flow cells.

Fig. 5c shows the comparison between OCEANS and NGS. All 16 somatic mutants called by NGS at above 5% VAF were also called by OCEANS, corresponding to a 100% OCEANS sensitivity relative to NGS. Of the 9584 NGS-negative loci, OCEANs called an additional 97 variants (Fig. 5c); thus, relative to NGS, OCEANS had a 99.0% specificity. By varying the VRF cutoff threshold, we can change the number of variant calls by OCEANS, generating a set of sensitivity/specificity tradeoffs which can be plotted as a receiver-operator characteristic (ROC) curve (Fig. 5d). The area under the ROC curve is 99.99%, indicating very high concordance between OCEANS and NGS when the OCEANS variant LoD is artificially weakened by setting higher VRF thresholds.

**TABLE 1:**
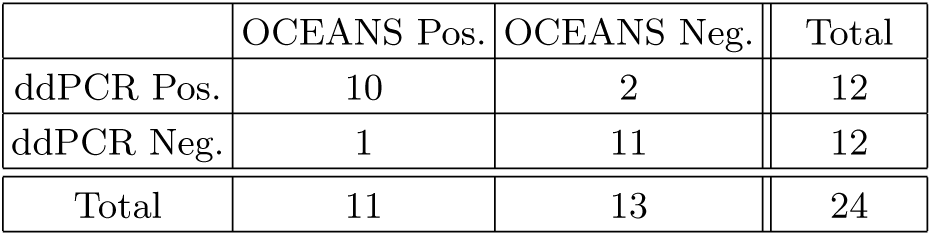
Summary of OCEANS and ddPCR comparison experiments for 6 FFPE samples in 4 select mutation loci (BRAF p. V600E, KRAS p. G13D, KRAS p. E62K, and MAP2K1 p. P124L). Other than one sample/mutation combination at 31% VAF, ddPCR showed VAFs ranging between 0.02% and 0.66% for the concordant samples. See Supplementary Section S6 for detailed results.

Importantly, we believe that many of the 97 discordant called variants based on a ≥ 20% VRF threshold and a Clair score ≥180 could be real mutations missed by NGS, based on our calibration experiments. In other words, relative to OCEANS results that we believe to be more accurate, standard NGS has a somatic mutation sensitivity of 14.2% and a specificity of 100%. To confirm our discordant OCEANS mutation calls, we further performed digital PCR on 6 FFPE samples at 4 mutation loci (BRAF p. V600E, KRAS p. G13D, KRAS p. E62K, and MAP2K1 p. P124L). Of these 24, 11 mutations were called positive by OCEANS, and 13 were called negative by OCEANS. OCEANS was concordant with ddPCR for 10 positive samples and 11 negative samples (Table 1, Supplementary Section S6).

It is important to note that concordant positives between OCEANS and ddPCR indicate the existence of a DNA variant in the sample, which may not necessarily reflect a mutation in the patient. Cytosine deamination is a well-documented type of DNA damage frequently observed in DNA extracted from FFPE. We applied an FFPE damage repair kit to the FFPE DNA before performing OCEANS library preparation, but do not necessarily expect that all cytosine deaminations are repaired or excised. In particular, any repair kit based on cleaving/repairing uracils formed through the deamination of standard cytosine would not be able to rectify deamination of methylcytosines into thymines.

Because each ddPCR mutation requires a separate reaction, the ddPCR results required 4 times more input DNA than OCEANS just to cover these 4 mutations. For analysis of clinical biopsy samples, OCEANS would have significantly higher clinical sensitivity due to being able to analyze all mutations in the panel from a single sample.

Next, we wished to characterize the reproducibility and robustness of the OCEANS panel on different types of nanopore sequencing instruments and flow cells. The Oxford Nanopore Flongle flow cell, in particular, is relatively inexpensive at $70, and can further reduce turnaround time relative to MinION by reducing the need for sample batching before sequencing. We performed the OCEANS panel on all 25 melanoma samples on the Flongle, and observed highly quantitatively similar VRFs as our results on the MinION.

### NSCLC and HCC OCEANS Panels

We next constructed two additional OCEANS panels: a 28-amplicon panel for non-small cell lung cancer (NSCLC) and an 11-amplicon panel for hepatocarcinoma (HCC) to show the generality of our approach. The NSCLC OCEANS panel covers roughly 1121 mutations in the COSMIC database across 13 genes (AKT1, ALK, BRAF, DDR2, EGFR, KRAS, NRAS, MAP2K1, MET, PIK3CA, PTEN, ROS1 and TP53, see Supplementary Section S4). DNA from 5 FF and 18 FFPE NSCLC tissue samples were sequenced using both OCEANS and NGS. Fig. 6ab show the comparison between OCEANS and NGS. 9 out of 11 somatic mutants called by NGS at above 5% VAF were also called by OCEANS. The two mutations that had a Clair score less than 180 were indel mutations, for which Clair has been observed to be less accurate [10, 25].

**FIG. 6:**
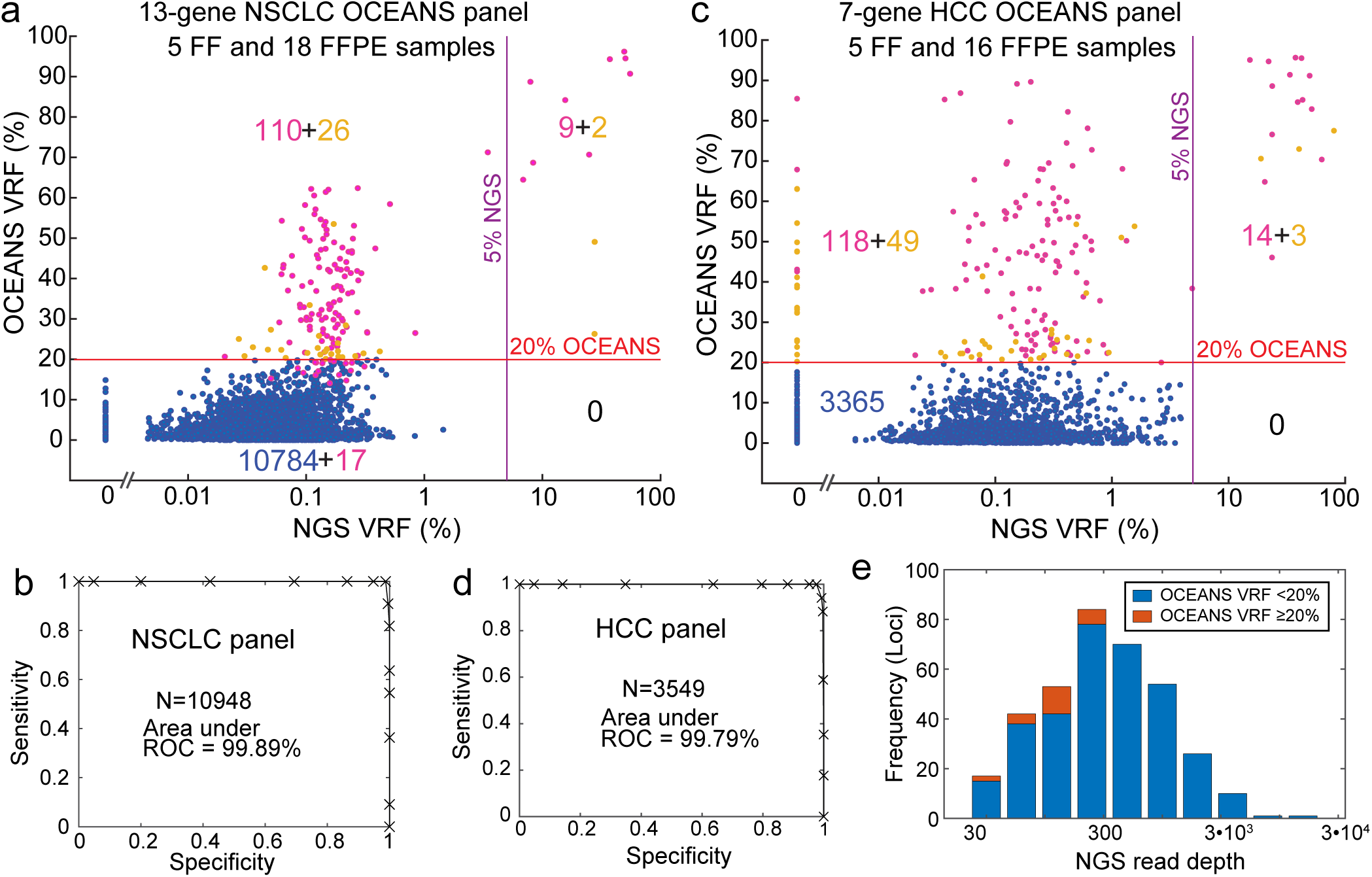
Validation of NSCLC and HCC OCEANS panels on clinical tumor tissue samples. **(a)** Summary of NSCLC panel results on 23 clinical samples (5 FF, 18 FFPE). Input DNA quantities ranged from 10 ng to 50 ng (Supplementary Section S5). The X-axis shows the VRF based on a standard NGS analysis, and the Y-axis shows the OCEANS VRF. The horizontal line shows the 20% VRF cutoff for OCEANS variant calls, and the vertical line shows the 5% VRF cutoff for NGS variant calls. The numbers in quadrants display the number of loci in each group; 110 of the 136 putative variants in the top-left quadrant also had Clair scores of above 180 (purple dots). 2 of the 11 NGS confirmed variants in the top-right quadrant had Clair scores below 180 (yellow dots); both variants were insertions that typically have lower Clair scores. **(b)** Receiver operator characteristic (ROC) curve for data in panel (a), based on changing the VRF cutoff for OCEANS. **(c)** Summary of HCC panel results on 21 clinical samples (5 FF, 16 FFPE). Input DNA quantities ranged from 10 ng to 50 ng (Supplementary Section S5). 118 of the 167 putative variants in the top-left quadrant also had Clair scores of above 180 (purple dots). 3 of the 17 NGS confirmed variants in the top-right quadrant had Clair scores below 180 (yellow dots); all 3 variants were in TERT amplicon within a homopolymer region that resulted in lower Clair scores (Supplementary Section S5). **(d)** ROC curve for data in panel (c), based on changing the VRF cutoff for OCEANS. **(e)** Panel (c) showed a significant number of loci wherein NGS results had 0% variant read frequency (VRF). Here, we show a histogram showing NGS read depth distribution for the 0% VRF loci, as well as the fraction of loci for each read depth group where OCEANS VRF was greater than 20%. Only loci with less than 300x NGS read depth showed discordance with OCEANS (mutation call based on 20% VRF).

The HCC OCEANS panel covers roughly 680 mutations across 7 genes (CTNNB1, ARID1A, AXIN, TERT, JAK1, PTEN and TP53, see Supplementary Section S4). DNA from 5 FF and 16 FFPE HCC tissue samples were sequenced using both OCEANS and NGS. Fig. 6cd show the comparison between OCEANS and NGS. 14 out of 17 somatic mutants called by NGS at above 5% VAF were also called by OCEANS. The 3 mutations not called by Clair were in the TERT amplicon within a homopolymer region (Supplementary Section S5). Higher NS error rates in homopolymer regions could be the reason for lower Clair scores despite the OCEANS VRF being >70% for these mutations. We observed 23 loci with OCEANS VRF greater than or equal to 20% for which the corresponding NGS VRF were 0%. We analyzed the NGS read depth for all loci with NGS VRF equal to 0% and their corresponding OCEANS VRF (Fig. 6e). All 23 loci with OCEANS VRF greater than or equal to 20% had NGS read depth of less than 300. Overall, both OCEANS panels had high concordance between OCEANS and NGS as shown by area under the ROC curve of 99.89% for the NSCLC panel and 99.79% for the HCC panel.

## Discussion

The rapid turnaround time and low instrument/consumables cost of nanopore sequencing render NS attractive for time-sensitive therapy selection and recurrence monitoring applications in oncology. The largest drawback of NS is its high intrinsic error rate. Although multi-nucleotide indels, copy number variations, and gene fusions can be detected at low VAFs by NS despite its error rate, previously the detection of single nucleotide somatic mutations with low VAFs was not possible by NS. The OCEANS method we present allows NS to detect single-base mutations from FFPE tissue-derived DNA with LoD of less than 1% with a 10 hour workflow, and positions NS to transition into clinical sequencing panels (Table 2). Within OCEANS, BDA enhances the confidence and sensitivity of somatic mutation calls, and SAL reduces the turnaround time by enhancing the throughput of NS.

**TABLE 2:** Comparison of sequencing methods.

|  | OCEANS<br>(MinION) | Standard NS<br>(MinION) | Standard NGS<br>(MiSeq) | NGS w/ UMI<br>(MiSeq) |
| --- | --- | --- | --- | --- |
| Samples / Flow cell | 24 | 1 | 24 | 2 |
| Flow cell price | \$900 | \$900 | \$1300 | \$1300 |
| Flow cell reusability | 10 | 10 | 1 | 1 |
| Sequencing price / sample | \$3.75 | \$90 | \$54.20 | \$650 |
| Turnaround time | 10 hr | 3 hr | 72+ hr | 72+ hr |
| Single-base VAF LoD | 0.05%-1% | 20+% | 2%-5% | 0.1%-0.5% |

For liquid tumors and for solid tumors where fresh/frozen tissue biopsy samples are available, the long read length of NS further enables accurate detection of large-scale DNA structural variations. Integrating OCEANS with existing NS experimental and bioinformatics methods for structural variant profiling would allow for the development of comprehensive oncology panels covering a broad range of structural alterations observed in leukemias and lymphomas. In contrast, the DNA from FFPE tissue samples and cell-free DNA from plasma are short and do not physically allow for direct long-read sequencing to identify large-scale alterations.

In this work, our bioinformatics pipelines make qualitative variant calls based on a combination of observed VRF and Clair score. In principle, the fold-enrichment could be calculated for each mutation in BDA [17], and the sample VAF could be inferred from VRF for OCEANS (Supplementary Section S4). In practice, however, the high NS error rates combined with the saturation of VRFs near 100% after BDA enrichment means that our quantitation dynamic range is relatively small. Integration of OCEANS with unique molecular identifier (UMI) barcodes [23, 24] will likely be necessary for accurate VAF quantitation on the NS platform.

## Data Availability

Custom bioinformatics software were written in python for NS reads process and are available upon request. Nanopore raw FAST5 and FASTQ data are available upon request. Summary of NS and NGS data, including number of reads mapped to wildtype and variant sequences in each amplicon, are included in the Supplementary Excel Table.

## Acknowledgements

This work was supported by NIH grant R01CA203964 to DYZ. The authors thank Jianyi Nie for editorial assistance. The authors thank Gang Bao for use of his group’s Bio-Rad ddPCR instrument. The authors thank Oxford Nanopore Technologies for providing MinION and Flongle flow cells. The authors thank Christina Hao for bioinformatics assistance.

## Author contributions

DT and DYZ conceived the project. DT designed and performed SAL feasibility and characterization experiments. DT and LYC designed OCEANS panels, with assistance from PS for BDA primer/blocker design. DT and SXC wrote bioinformatics software for inferring VRF from OCEANS and NGS FASTQ data. LK and MJB provided clinical samples. PJ and DJT performed independent validation of OCEANS panels at Oxford Nanopore Technologies, and wrote bioinformatic deconcatenation software. DT and DYZ wrote the manuscript with input from all authors.

