## Supplementary info for "Oncogene Concatenated Enriched Amplicon Nanopore Sequencing for Rapid, Accurate, and Affordable Somatic Mutation Detection"

(Dated: November 10, 2020)

S0. Materials and Methods 1
S1. Effect of DNA Length on NS Throughput 3
S2. SAL Design and Performance 5
S3. Bioinformatic Analysis 8
S4. Analytical Validation Experiments with Synthetic DNA 12
S5. Mutations Detected in Melanoma Clinical Samples and NGS Comparison 15
S6. ddPCR Comparison 18

#### Section S0: Materials and Methods

**Oligonucleotides.** Primers and Blockers were purchased from Integrated DNA Technologies, as well as gBlocks that serve as positive controls. All DNA oligos were purchased standard desalted and dissolved in 1x TE buffer (10 mM Tris-HCl, 1 mM EDTA, pH 8). Dilutions of gBlock DNA were done in 1x TE buffer with 0.2% Tween 20 (Sigma) and 100 ng/ $\mu$l Carrier RNA(poly A)(Qiagen, Catalog No.1017647).

**Repository DNA samples.** Human gDNA samples (NA18537 and NA18562) were purchased from Coriell Biorepository. BRAF V600E reference standard (HD238) and Myeloid DNA reference standard (HD829) were purchased from Horizon Discovery. Seven melanoma fresh frozen patient tissue samples were purchased from OriGene. Twenty-one melanoma FFPE patient tissue samples were obtained deidentified from MD Anderson Cancer Center. Five HCC and five NSCLC fresh frozen patient tissue samples were purchased from OriGene. Fourteen HCC FFPE patient tissue samples were purchased from US Biolab and two HCC FFPE patient tissue samples were purchased from OriGene. Eighteen NSCLC FFPE patient tissue samples were purchased from OriGene. DNA from fresh frozen samples were extracted using QIAamp DNA Mini Kit (Catalog No. 51304). DNA from FFPE samples were extracted using Qiagen GeneRead DNA FFPE kit (Catalog No.180134) and repaired using NEBNext FFPE DNA repair mix (NEB #M6630L).

**Stochastic Amplicon Ligation (SAL).** BsaI restriction sites and complementary overhangs for assembly were appended to DNA by PCR using SAL adapter primers and Phusion Hot Start Flex DNA polymerase (NEB M0535S). For single-plex reactions, the following thermocycling protocol was used: 98°C- 30s; (98°C- 20s, 63°C- 30s, 72°C- 30s)x 3; (98°C- 20s, 72°C- 1 min)x n; 72°C- 5 min. For multiplex panels, the following thermocycling protocol was used: 98°C- 30s; (98°C- 20s, 63°C- 2 min, 72°C- 2 min)x 3; (98°C- 20s, 72°C- 3 min)x n; 72°C- 5 min. The number of PCR cycles (n) was empirically determined based on the amount and nature of input DNA. Amplicons from PCR were purified by column purification with Monarch PCR & DNA Cleanup Kit (NEB #T1030). DNA was quantitated by Invitrogen Qubit dsDNA HS (High Sensitivity) Assay Kit (Catalog number: Q32851).NEB BsaI-HFv2 kit (E1601S) was used for amplicon assembly. A 20 $\mu$l reaction was set up containing 400-500 ng of DNA and 2$\mu$l golden gate enzyme mix in 1x T4 DNA ligase reaction buffer. Reaction was cycled for 30 times, with each cycle containing an incubation at 37 °C for 2 min and at 16 °C for 2 min. The assembled DNA was size selected twice using 0.4x Agencourt AMPure XP beads (Beckman Coulter A63881).

**Oncogene Concatenated Enriched Amplicon Nanopore Sequencing (OCEANS).** 10 ng to 50 ng of human genomic DNA sample was mixed with primers and blockers, and subject to PCR using Phusion Hot Start Flex DNA polymerase. For single-plex tests, 400 nM of the forward and reverse primer and 4 $\mu$M blocker were used. For the 7-plex AML panel, 75 nM of each forward and reverse primer and 750 nM of each blocker were used. For the 15-plex melanoma panel and 11-plex HCC panel, 50 nM of each forward and reverse primer and 500 nM of each blocker were used. For the 28-plex NSCLC panel, 15 nM of each forward and reverse primer and 150 nM of each blocker were used. For single-plex, the following thermocycling protocol was used: 98°C- 30s; (98°C- 10s, 63°C- 30s, 72°C- 30s) x23; 72°C- 5 min. For AML and Melanoma multi-plex panels the following thermocycling protocol was used: 98°C- 30s; (98°C- 20s, 63°C- 2 min, 72°C- 2 min) x23; 72°C- 5 min. For HCC and NSCLC multi-plex panels, the following thermocycling protocol was used: 98°C- 30s; (98°C- 10s, 63°C- 5 min, 72°C- 30s) x23; 72°C- 5 min.

The amplicons were then purified by column purification, and used as input for PCR using SAL adapter primers and subject to SAL as described above. Assembled DNA was used for library preparation using Ligation sequencing kit (SQK-LSK109) following the protocol provided by ONT. Briefly, 50ng - 200 ng of assembled DNA was end-repaired and dA tailed using NEBNext Ultra$^{TM}$ II End Repair/dA-Tailing Module (E7546) and purified using 1x AMPure XP beads. Native barcoding kit (EXP-NBD104 or EXP-NBD114) was used for barcoding. 40-100 fmol pooled barcoded DNA was used for adapter ligation. The ligation reaction was purified using 0.5x AMPure XP beads. The beads were washed using S Fragment Buffer (SFB) and eluted in Elution Buffer (EB). The eluate was quantified by Qubit and then loaded on to Minion R9.4.1 flow cells, and sequenced for 40 min. For Flongle flow cells, 20-50 fmol pooled barcoded DNA was used for adapter ligation, and sequencing was run for 8-10 h.

**Next Generation Sequencing (NGS) verification of clinical samples on Illumina.** 10 ng to 50 ng of genomic DNA extracted from FFPE samples were used as input, for multiplex PCR amplicon sequencing. The following thermocycling protocol was used: 98°C- 30s; (98°C- 20s, 63°C- 2 min, 72°C- 2 min)x 15; 72°C- 5 min. The amplicons were purified by column purification. NEBNext ultra II DNA library prep kit for Illumina (NEB # E7645S) was used for library preparation following the kit protocol. NEBNext Multiplex Oligos for Illumina (Dual Index Primers Set 1) (NEB #E7600S) were used for index PCR. Sequencing was done on Illumina MiSeq using the

V2 kit. Each sample was sequenced to at least 10,000x coverage. Libraries were checked by running on Bioanalyzer before sequencing.

**ddPCR quantitation protocol.** ddPCR assays were performed on QX200 Droplet Digital PCR system (Bio-Rad) using BRAF V600E (Bio-Rad dHsaMDV2010027), BRAF V600K (Bio-Rad dHsaMDV2010035), KRAS p.E62K (Bio-Rad dHsaMDS453364969), KRAS p.G13D c.38G>A (Bio-Rad dHsaMDV2510598), and MAP2K1 p.P124L (Bio-Rad dHsaMDS897199134) mutation detection kits according to kit protocols.

**NS Bioinformatic analysis**. NS reads were basecalled using MinKNOW 19.12.5 (fast basecalling model). Barcoded NS fastq reads were demultiplexed using EPI2ME software (Oxford Nanopore Technologies). Typically, 10,000 NS reads were used per sample for analysis. The SAL reads were deconcatenated using a custom python script (Available upon request) and the deconcatenated reads were aligned to human reference genome (GRCh38) using minimap2 aligner (https://github.com/lh3/minimap2) to generate a bam file. IGVtools (http://software.broadinstitute.org/software/igv/download) was used to extract the number of A, C, G & T basecalled nucleotides, insertions and deletions at each position of the amplicon using the basecount command in IGV-command-line tools. The VRF at each nucleotide position was calculated as the highest frequency single-base change at that position.

Bioinformatic workflow for variant calling using Clair variant caller is summarized in Fig. S3-1. Clair v1 (https://github.com/aquaskyline/clair) was used for variant calling. First, the bam file generated using minimap2 was down-sampled to <150x coverage for each amplicon using samtools view command. The subsampled bam file was then used to call variants using callVarBam submodule and ont r94-flipflop model. Allele frequency threshold of 0.2 and minimum coverage of 50x were used. Variant calls were filtered for score >180.

**NGS Bioinformatic analysis**. NGS reads were trimmed of adapter sequences and aligned to reference amplicon sequence using short read (sr) function of minimap2 aligner. IGVtools (http://software.broadinstitute.org/software/igv/download) was used to extract the number of A, C, G & T basecalled nucleotides, insertions and deletions at each position of the amplicon using the basecount command in IGV-command-line tools. The VRF at each nucleotide position in the enrichment region (nucleotide positions covered by the blocker in BDA) was calculated as the highest frequency single-base change at that position. Mutations in the enrichment region with VRF >5% and minimum coverage of 80x were considered true positives.

#### Section S1. Effect of DNA Length on NS Throughput

**NS of 160 bp vs 10 kb fragmented human genomic DNA**. Fig. S1-1 shows gel images of human genomic DNA fragmented to mean length of 160 bp or 10 kb. NS read statistics of the two libraries are shown in Fig. S1-2 and Fig. S1-3. The mean read length of 160 bp fragmented library was 265 bp, due to the attachment of barcodes and sequencing adapter prior to NS. The 10kb DNA library had a mean read length of 3.8 kb. The Q-score of 10 kb library was higher than that of the 265 bp library.

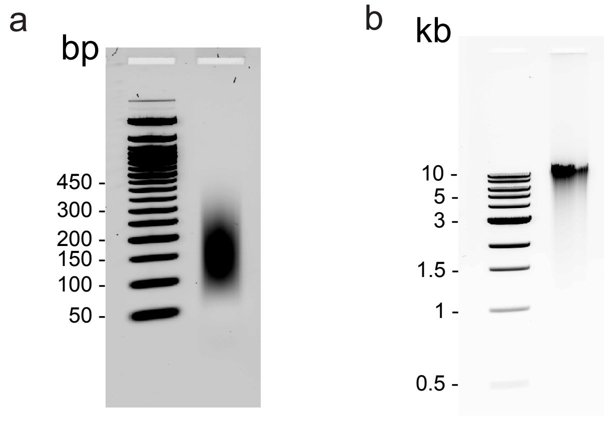


FIG. S1-1: Size verification of fragmented human genomic DNA. **(a)** 160 bp library run on a 2% agarose gel. **(b)** 10 kb library run on a 0.5% agarose gel.

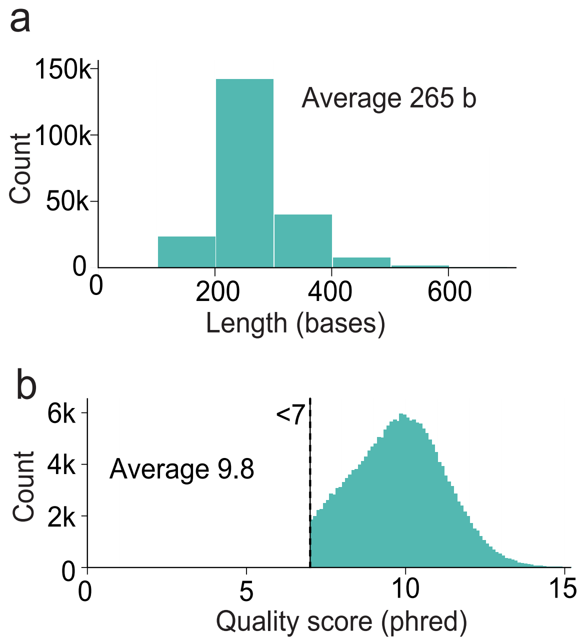


FIG. S1-2: NS run performance of 160 bp fragmented library. **(a)** Length distribution of NS reads. **(b)** Q-score distribution of NS reads.

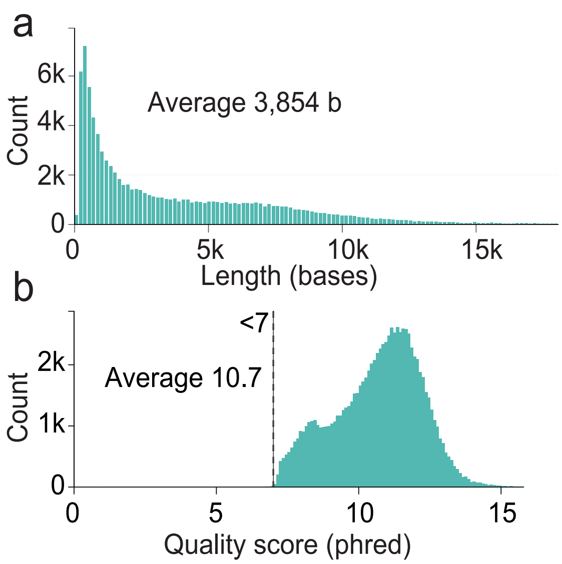


FIG. S1-3: NS run performance of 10 kb fragmented library. **(a)** Length distribution of NS reads. **(b)** Q-score distribution of NS reads.

### Section S2: SAL Design and Performance

Fig. S2-1 shows the design of SAL adapter sequences. Fig. S2-2 shows agarose gel electrophoresis analysis of SAL concatemer products before and after exonuclease digestion. The remaining bands after digestion are likely circular concatemer products. NS throughput for original amplicons vs. SAL products are shown in Fig. S2-3. Fig. S2-4 shows the individual NS amplicon traces for 0% and 5% VAF samples shown in Fig. 2e.

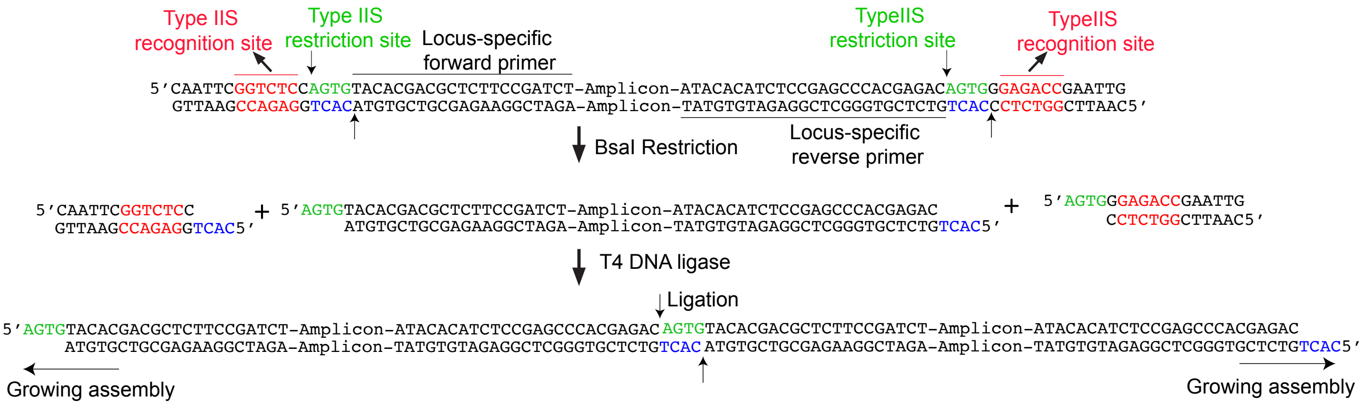


FIG. S2-1. Schematic of SAL design. Adapter sequences containing type IIS recognition sequence (red) followed by restriction sites are attached to the ends of amplicons by PCR. After cleave by the type IIS restriction enzyme BsaI, complementary 4 base overhangs (green and blue) are generated. During the ligation step, two amplicons hybridize through their complementary ends are ligated. The DNA assembly grows at both ends by cycling between restriction and ligation steps.

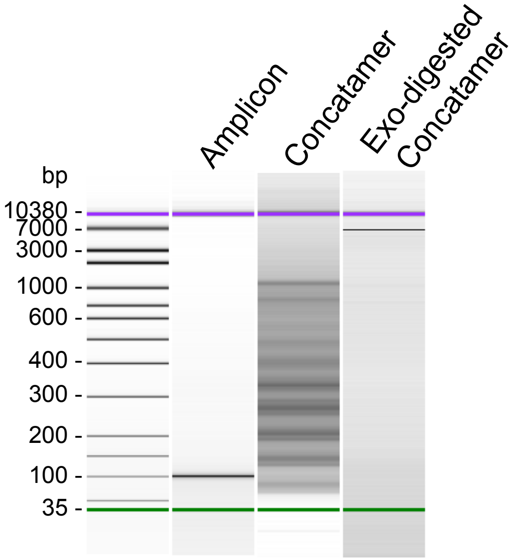


FIG. S2-2. Exonuclease treatment of SAL concatemers. 25 ng of SAL assembled concatemers of 100 bp DNA were treated with 10U of T7 exonuclease and 10U of Exonuclease VII to digest linear dsDNA.

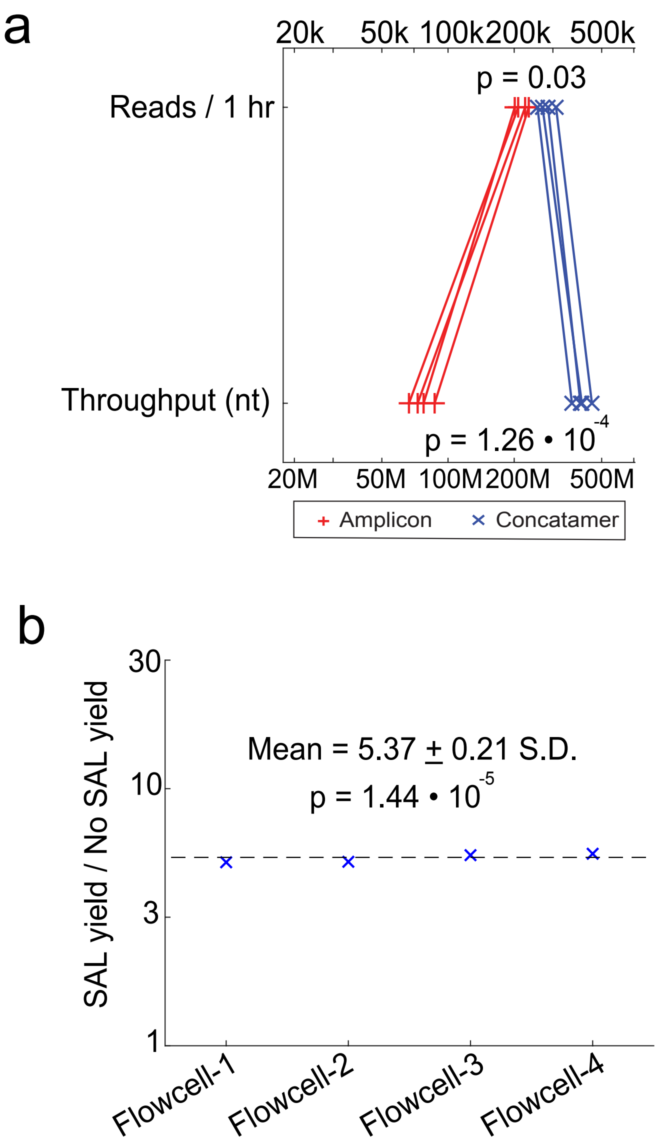


FIG. S2-3. NS throughput when sequencing amplicons vs. SAL concatemers. **(a)** NS reads and throughput for amplicons vs. concatamers. Here, PCR amplicons of size 180bp were assembled by SAL. The amplicon without SAL and after SAL were sequenced on four new flow cells for 1 hour each. In two flow cells, the amplicon library was sequenced first, the flow cell was then washed and loaded with the SAL library. The order of loading was reversed for the other two flow cells. $p$-value was calculated using two-way Student's t-test. **(b)** Ratio of yield of direct amplicon library to SAL library on each flow cell plotted on log scale, with $p$ value calculated via a one-way Student's t-test.

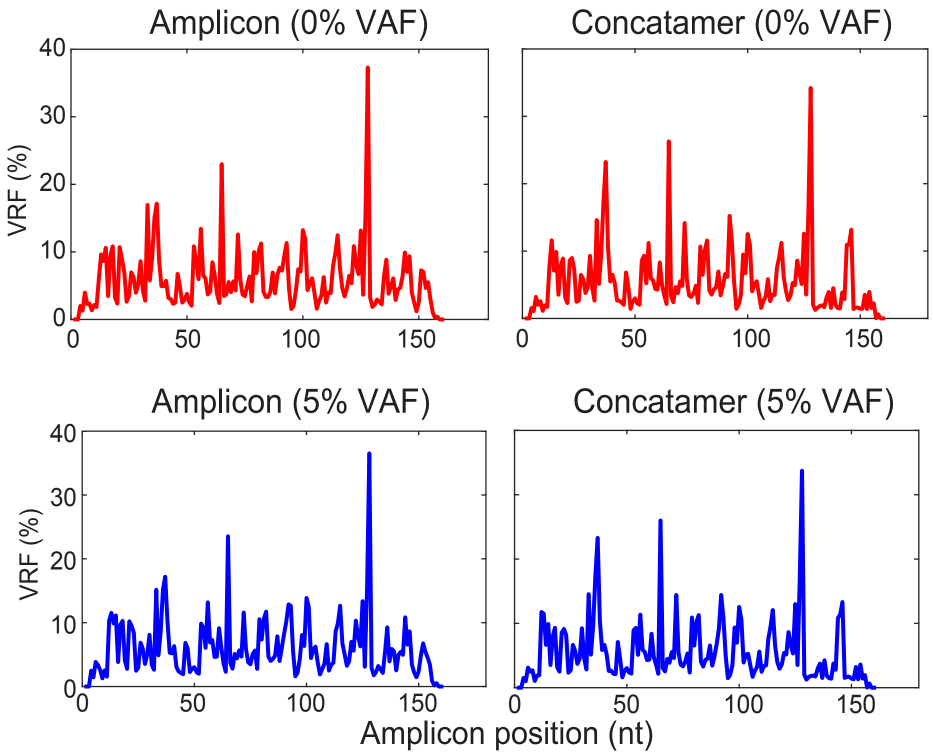


FIG. S2-4. NS amplicon traces with and without SAL. Top panels show amplicon traces for 0% VAF sample. Bottom panels show amplicon traces for 5% VAF sample. VRF of NS reads at each location that corresponds to the highest frequency single-base changes at that position. ΔVRF in Fig. 2e was calculated by subtracting VRF of 0% VAF sample from 5% VAF sample.

#### Section S3: Bioinformatic Analysis

**Summary of bioinformatic workflow.** NS on MinION was run for 30 min to 1 hour depending on the number of barcoded samples. Typically, 350k-500k reads were obtained in the first one hour of NS. Base-called reads were demultiplexed using EPI2ME software (Oxford Nanopore Technologies). Depending on the total number of reads obtained per sample, reads were subsampled randomly to approximately 10,000 reads per sample for analysis. The NS reads were then deconcatenated using a custom python script.

The python script used minimap2 (github.com/lh3/minimap2) to map individual wildtype amplicon reference sequences to the concatenated reads and extract the mapped monomer sequences from the concatenated read. This step also removed any off-target amplicon sequences from the concatenated reads. Fig. S3-1 shows a sample concatenated read.

The mapping efficiency for finding monomer sequences within concatenated reads was calculated as mapped nucleotide fraction (MNF), the fraction of concatemer reads that mapped to an amplicon of interest (Fig. S3-1). Manual analysis of concatenated reads showed that minimap2 missed some monomers (Fig. S3-1). Tables S3-1 and S3-2 shows the number of reads analyzed for each sample and the MNF values. MNF was higher for FFPE samples, which could be due to the fragmented nature of DNA that resulted in reduced amplification of off-target sequences. Sub-sampling different sets of approximately 10,000 reads resulted in same variant calls with similar % VRF values (Table S3-3).

The mapped on-target sequences obtained after deconcatenation were then analyzed in two ways for variant calling. First, the sequences were aligned to amplicon reference sequences using minimap2 aligner to generate a BAM alignment file. IGVtools was used on the BAM file to extract the number of A, C, G, and basecalled nucleotides, insertions and deletions at each position using the basecount command. The VRF at each nucleotide position was calculated as the highest frequency single-base change at that position. Positions with VRF ¿20% were flagged as potential variants.

Second, the sequences were aligned to human reference genome (GRCh38) using minimap2 aligner to generate a BAM alignment file. The BAM file was down-sampled to <150x coverage for each amplicon using samtools view command. The downsampled BAM file was then used with Clair to call variants. Variant calls with score > 180 were flagged as potential variants. The bioinformatic workflow for variant calling using Clair variant caller is summarized in Fig. S3-2. Potential variants with both VRF> 20% and CLAIR score > 180 were definitively called as variants.

#### Sample SAL concatenated read (2925 nt)

CAATTCGGTCTCCCACTAGATGATGGGCTCCCGGAAGACAGTCCCCCAGGATGTTCGGATAGTTCCATTGGGACTTTCCACATCTTCT CACTCATCTGCAAAAACATCCCACGCCCC
TAGTCCCTGGCTGGACCAAGCCCATCACCATTGGCAGGCACGCCCATGGCGACCAACACTGTGTTGAGATGGACCCCCTATTTATGGATTTATTTGTATTTGCCTTTAGCTAAATGTG
TGTAAATACAGTTATACATATATGCATTCTCAATTTCATACCTTGCTTAATGAGGTGTAGATACCCAAAGATAAAGAATAAAAACACATACAAGTTGGAAATTTCTGGGCCATGAAAAA
AAAACATGCAAATCACATTATTGCCAACATGACTTGCCAGTCCCATAAGCATGACAACCTATGATGATAGGTTTACCCATCCTCAAAAGCCACTCATCTGCAAAAACATCCCACGCCT
AGTCTAAGCTGCGTTGGCCCCATCACCGGTGGCAGGCACGCCCATGGCGACCAGCACTAGATGATGGGCTCCCGAAGTAGTCCCCCAGGGGATGTTGAATGGTTCCATTGGGACT
TTCACATCTTCTCACTAGTTAGTTTTCACTACTACAAGTTAAAATGAATTTAAATAGTTTCTTTCTCCTCCAACTAATAGTGTATTCCACAGAGACAGCAGCCAGAAATATCCTCCTTAC
TCATGGTCGGATCACAAGATTTGTGATTTTGGTCTATCAGACACATCAAGAATGATTCTAATTATGTGGATTTAAAGGACACTGAATGGGCTCCCAGAAGACAGTCCCCCAGGATGTT
CCAGATAGTTCCGTGGGACTTTTCCACATCTTCTCACTAGATGATGGGCTCCCGGAAGACAGTCCCCCAGGATGTTCAGATAGTTCCATTGGGACTTTTCCACATCTTCTCACTTGTG
TTGAGATGGACAACCTATTTGTAAAGTTTATTTGTATTTGCCTTTAGCTAAATGTGTGTAAATATATACAGTTATACATATGCATTTCTCAATTTCATACCTTGCTTAATAGATTGTAGATAC
CAAAGATAAGAATAAAACACATACAAGTTGGAAATTTCTGGGCCATAGAAAAAAAAAAAAAAACATGCAAAATCACATTATTGCCAACATGACTTGCTTGATCCCCATAAGCATGACG
ACCTATGATGATAGGTTTTACCCATCACTCACAAGCCACTAGATGGTGGGCTCCCGGTGGCATTTCTTGCCCGACCTGAGGATGTACCCGCCACGCCTGCAGGACTGACCTCTAGGT
GGGCAAGCCGAGGCACAAGAGGGCGCGCTCTGGCGGAGTCAGCCCTCTTGTGAATTTGGGCTGGGAGACCAGCCAGGCCTAACAGGCAAGGCTGGGTCAGGCGAGGTCCTGG
AGCCCACTTCGGTGTAGAGTGTGCAAAATGTGAACAGGCCTGCCCAGGATAGCTCCTGCATTAGCGCTGGTGCCTTACGAGCGGCCAGTCAGATTTTTATTTGGCACCACTACAGA
GAGACCCAGGAGAGTCTCTTTAAGAAAATAGTTTAAACCACTAGATGATGGGCTCCCGGAAGACAGTCCCCCAGGATGTTCGGATAGTTCCGTGGGACTTTTCCATCTTCTCACTCC
CGCCAGGAACGTGCTTGTCACCCACGGGAAAGTGGTGAGAATATGTGACTTTGGATTAGCTCGAGTTATCATGAGTGATTCCAACTATGTTGTCAGGGCACTCATCTGCAAAAACAT
CCACACTAGTCCCTAGCTGGACCAAGCCCATCACCATTGGCAGGCACGCCCATGGCGACCAGCACTAGATGGGCTCCGGAAGACTGATCCCCAGGATGTTCCGGATAGTTCCATTG
GGACTTTCCACATCTTCTCACTCCGCCAAGGAACGTGCCTTGTCACCCACGGGAAAGTGGTGAAGATATGTGACTTTGGTTGGCTCGAGTTATCATGAGTGATTCCAACTATGTTGT
CAGGGCACTAGATGATGGGCTCCCCGGAAGACAGTCCCCAGGATGTTCCAGATAGTTCCATTGGGACTTTTCATCTTCTCACTAGATGATGGGCTCCCGGAAGACAGTCCCCCAGG
ATGTTCCAGATAGTTCCATTGGGACTTTTCATCTTCTCACTAGAAGATTTCCTTGGAACTAAGCAGGCGTCAGAGGAGTTGGTGGGTGTGAGTGCCCTGTCCCTGCACTTCGGGTGG
CTGCTGGTCCTCAGGTCCTGCTGTGTGGTTAGACGGCTTCCGGGCAGCCTGGTCTGGCCAACACTCACCTACCCTCTCTGCCTTTTCTCCCCAGAATTGTTGGTTTCCCAGTCCACTA
TACTGACGTCTCCAACATGAGCCGCTTGGCGAGGCAGAGACTGCTCACTTTAAAGTGTTTGGAATTAAATTACATCTAATTATAAAAATTTTCTTGGAGTCATATCTTTATCTAGAGTTA
ACTCTCTGAGTGGTAGAATGAAAAAAACAGATGTTGAACTATGCAAGAGACATTTGAATTTATTGATGTCTATGAAGTGTTGTGGTTCCTTAGCCACATTTCTTTTTTTCAGGCTATTC
AAGATCTCTGCATGGCAGTGGAGGAAGTCTCTTTAAAATAGTTTAAACCACTTTAAAGTGTTTGGACTAAATTACAAATACGAGTATAGGGTACTGCTTGGAGTCATATCTTTATCTAG
AGTTAACTCTCTGGTGGTAGAATGAAAGTAGATATTGAACTATGCAAAGAGACATTTAATTTGATGTCTGTGAGTGTCATTCTGTTCATCTCCATCATGGCGGTGGAGGAAGTCTCTTT
AAGAAAATAGTTTAAACCACTAGATGATGGGCTCCCGGCCTCCTCTCCATTCAATGCCTGCCCCAACTCCCTGAGCTCTAGCTCCGCTGGTCTCCTCCGAGG

Amplicon monomers: DNMT3A KIT NPM1 IDH1 IDH2-140 IDH2-172 FLT3 Off-target

$$\textbf{Mapped nucleotide fraction (MNF)} = \frac{\textbf{[Length of Mapped (Underlined) reads]}}{\textbf{[Length of concatamer]}} \times 100\%$$

$$= \frac{1711\ \text{nt}}{2925\ \text{nt}} \times 100\% = 58.5\%$$

FIG. S3-1. Sample SAL concatenated read obtained on the OCEANS AML panel. Different colors show the 7 different amplicon sequences of the AML 7-plex panel. All the amplicons of the panel happen to be present in this concatenated read. Two off-target (brown) sequences are also present. CACT sequence (black) is the 4 nucleotide overhang sequence used in the SAL design. Underlined sequences are those that were mapped by minimap2 and deconcatenated by the custom python code. The deconcatenated MNF was calculated to be 58.5%.

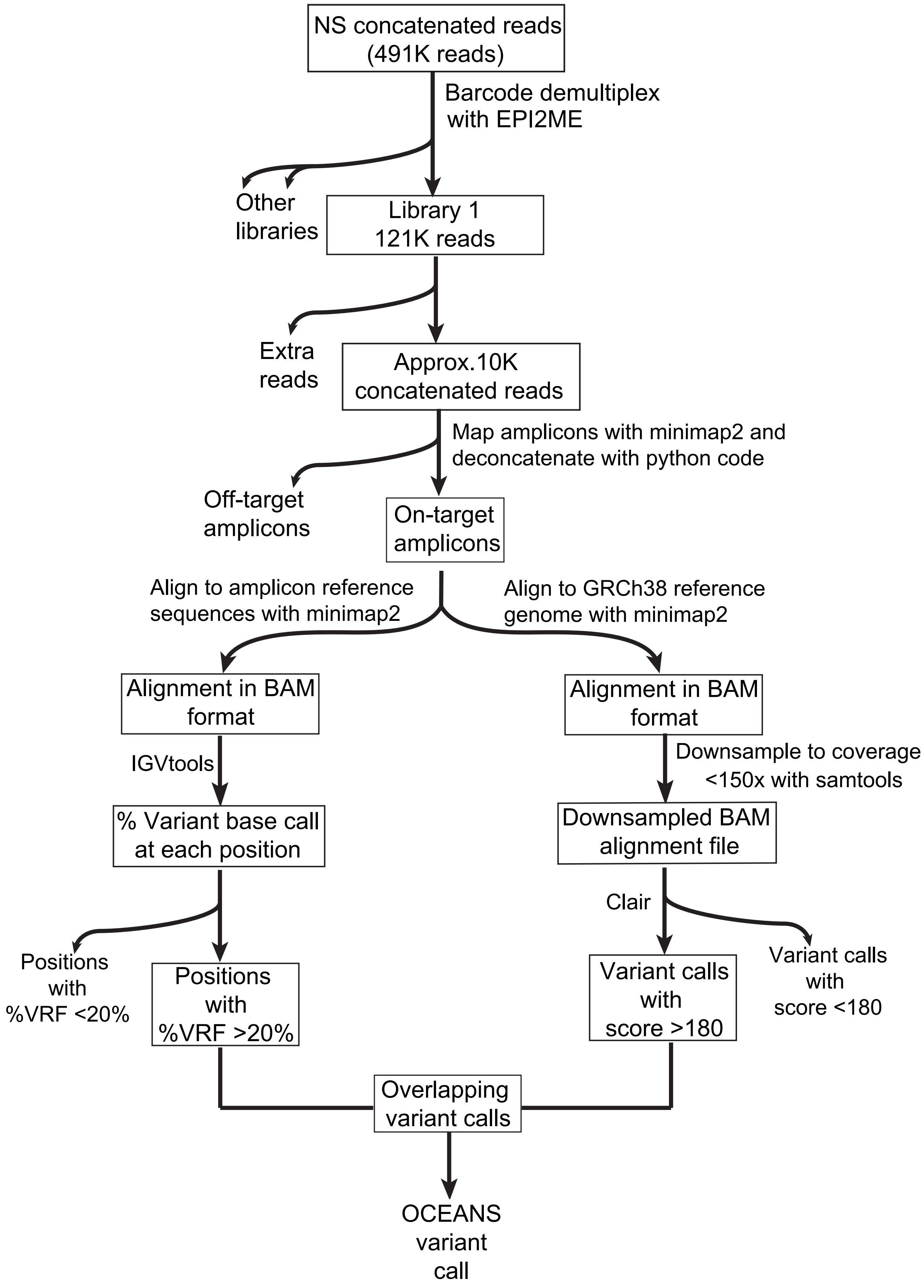


FIG. S3-2. Schematic of bioinformatic workflow for OCEANS variant calling.

| Sample | NS reads | NS throughput (Mb) | Down-sampled reads | MNF |
|---|---|---|---|---|
| Synthetic 0% | 40371 | 38.84 | 9948 | 22.66% |
| Synthetic 0.05% | 47480 | 40.24 | 10548 | 32.07% |
| Synthetic 0.1% | 46662 | 39.36 | 10361 | 35.05% |
| Synthetic 0.2% | 48400 | 39.20 | 10916 | 41.17% |
| Synthetic 0.5% | 49076 | 40.81 | 10920 | 45.14% |
| Synthetic 1.0% | 38956 | 36.74 | 9777 | 48.71% |
| Horizon HD238 | 103621 | 89.98 | 11366 | 33.38% |
| Clinical FF2 | 106152 | 102.89 | 8305 | 33.25% |
| Clinical FF3 | 100151 | 103.39 | 8321 | 29.37% |
| Clinical FF20 | 101103 | 95.33 | 8601 | 41.76% |
| Clinical FF26 | 99803 | 91.88 | 7951 | 38.05% |
| Clinical FF52 | 110598 | 103.63 | 8762 | 50.48% |
| Clinical FF61 | 121686 | 106.37 | 9049 | 29.53% |
| Clinical FF172 | 134055 | 104.32 | 10165 | 38.43% |
| Clinical FFPE3 | 167541 | 119.61 | 8965 | 54.34% |
| Clinical FFPE4 | 131576 | 104.35 | 9066 | 61.42% |
| Clinical FFPE5 | 124667 | 104.31 | 11127 | 63.51% |
| Clinical FFPE7 | 152421 | 115.09 | 9986 | 60.51% |
| Clinical FFPE8 | 127153 | 96.82 | 9429 | 59.97% |
| Clinical FFPE10 | 115243 | 87.26 | 10381 | 62.78% |
| Clinical FFPE12 | 141370 | 110.07 | 10879 | 57.78% |
| Clinical FFPE13 | 130721 | 112.89 | 9565 | 51.72% |
| Clinical FFPE14 | 157380 | 119.88 | 9137 | 53.53% |
| Clinical FFPE15 | 118950 | 105.75 | 9055 | 60.53% |
| Clinical FFPE17 | 179188 | 114.15 | 9999 | 49.36% |
| Clinical FFPE18 | 123029 | 92.40 | 10013 | 65.05% |
| Clinical FFPE19 | 160886 | 120.33 | 10643 | 58.53% |
| Clinical FFPE20 | 178399 | 133.02 | 9844 | 58.3% |
| Clinical FFPE21 | 135317 | 108.29 | 10147 | 58.42% |
| Clinical FFPE23 | 118125 | 99.96 | 8095 | 54.16% |
| Clinical FFPE24 | 118941 | 97.34 | 9592 | 56.26% |
| Clinical FFPE25 | 120325 | 108.32 | 9886 | 54.7% |

TABLE S3-1. Number of reads obtained and sampled for OCEANS melanoma panel, and the MNF for each sample.

| | DNMT3A | IDH1 | FLT3 | IDH2 | NPM1 |
|---|---|---|---|---|---|
| Subsample 1 | 47.79 | 95.08 | 92.97 | 58.04 | 46.36 |
| Subsample 2 | 51.72 | 91.23 | 91.67 | 59.62 | 47.45 |
| Subsample 3 | 45.05 | 93.69 | 89.55 | 60.66 | 45.27 |
| Subsample 4 | 42.11 | 89.92 | 92.31 | 53.85 | 41.77 |
| Subsample 5 | 53.76 | 93.81 | 89.04 | 53.85 | 39.88 |
| Subsample 6 | 50.00 | 90.35 | 93.18 | 46.67 | 42.86 |
| Subsample 7 | 46.25 | 95.38 | 91.57 | 47.54 | 44.94 |
| Subsample 8 | 61.84 | 89.23 | 95.89 | 51.06 | 45.51 |
| Subsample 9 | 42.22 | 93.58 | 95.79 | 56.60 | 41.90 |
| Subsample 10 | 47.78 | 93.86 | 92.00 | 57.69 | 41.13 |
| Subsample 11 | 41.56 | 92.54 | 93.24 | 49.09 | 41.21 |
| Subsample 12 | 43.68 | 91.82 | 91.18 | 56.25 | 42.86 |
| Subsample 13 | 55.41 | 93.97 | 84.62 | 40.35 | 40.48 |
| Subsample 14 | 48.86 | 91.04 | 89.53 | 63.16 | 37.50 |
| Subsample 15 | 46.43 | 82.98 | 92.68 | 54.84 | 37.36 |
| Mean | 48.30 | 91.90 | 91.68 | 53.95 | 42.43 |
| Std. | 5.60 | 3.10 | 2.79 | 6.08 | 3.04 |

TABLE S3-2. Variation in VRF (%) for different subsamples of 10,000 concatenated reads for the Horizon myeloid reference (HD829).

#### Section S4: Analytical Validation Experiments with Synthetic DNA

**Analytical validation experiments using synthetic DNA spike-in samples.** Multi-gene OCEANS panels were calibrated by a spiking synthetic mutation-bearing DNA oligonucleotide (gBlock) into the human NA18562 gDNA. The 10% VAF positive control sample had its VAF confirmed by NGS, and was diluted with NA18562 gDNA to prepare positive control samples with VAF ranging between 1% and 0.05%. For 1% and 0.5% VAF samples, 50 ng input DNA was used for OCEANS panels, corresponding to 15,000 haploid copies. For 0.2%, 0.1%, and 0.05% VAF samples, 100 ng input DNA was used for OCEANS panels.

OCEANS reads were deconcatenated, aligned to human reference genome (GRCh38), and then both the Clair score and the Variant Read Fraction (VRF) were calculated for each mutation. A summary of the variant calls made by Clair for melanoma OCEANS panel's calibration runs is shown in Fig. S4-1. Chromosome positions covered by the OCEANS panels are shown in Table S4-1 to Table S4-4. Spike-in mutations tested and their computed median enrichment fold (EF) values for AML and melanoma panel are shown in Table S4-5 and Table S4-6.

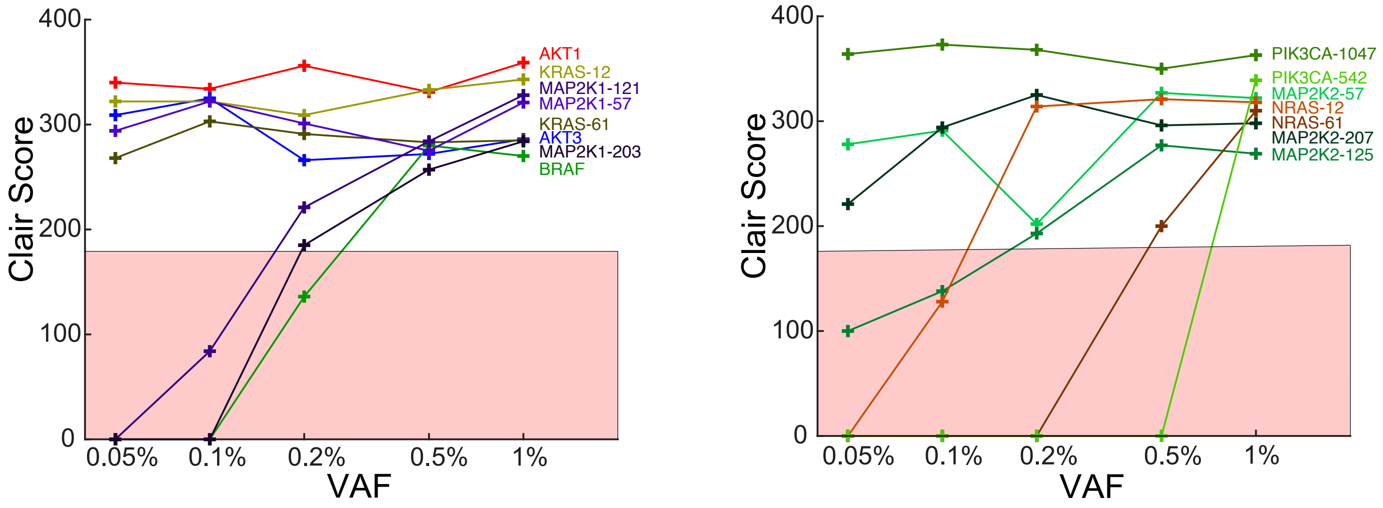


FIG. S4-1. Summary of Clair score for OCEANS melanoma panel. Mutations with Clair score > 180 are considered high confidence mutations.

**Enrichment fold (EF) calculation.** Enrichment fold was calculated as described previously in ref. [1]. Briefly, EF was calculated from the variant read fraction (VRF) observed from NS after OCEANS and the variant allele fraction (VAF) in the original sample based on the formula:

$$EF = \frac{(\frac{1-VAF}{VAF})}{(\frac{1-VRF}{VRF})}$$

For the AML and melanoma panels, EF was calculated for each plex by spiking synthetic DNA bearing a mutation into NA18562 gDNA shown in Fig. 4a and Fig. 4d. Median EF was calculated from EF values in the linear range from 0.05% to 1% VAF for each plex. Given a EF value for a particular mutation, the original VAF of the mutation in the sample can, in principle, be calculated from the VRF based on the formula $VAF = \frac{VRF}{(EF-1)(1-VRF)+1}$. However, due to the relatively high variation in observed VRF due to nanopore sequencing's intrinsic error rate, the effective dynamic range of quantitation is small, and VAF estimations are accurate only when the observed VRF is between 10% and 90%. For example, given an EF value of 1000, 99% VRF and 95% VRF correspond to VAFs of 10% and 2%. At 85% vs. 80% VRF, the VAFs correspond to 0.56% and 0.40%, which is a relatively smaller difference.


[1] Song, P., Chen, S. X., Yan, Y. H., Pinto, A., Cheng, L. Y., Dai, P., Patel, A. A., & Zhang, D. Y. Detecting and Quantitating Low Fraction DNA Variants with Low-Depth Sequencing. BioRxiv, 2020.04.26.061747 (2020).

| Gene | Enrichment Region (GRCh38) |
|---|---|
| FLT3 | Chr13: 28,018,487-28,018,513 |
| DNMT3A | Chr2: 25,234,363-25,234,383 |
| IDH1 | Chr2: 208,248,383-208,248,413 |
| KIT | Chr4: 54,733,139-54,733,168 |
| NPM1 | Chr5: 171,410,531-171,410,557 |
| IDH2 | Chr15: 90,088,694-90,088,719 |
| IDH2 | Chr15: 90,088,599-90,088,623 |

TABLE S4-1. Chromosome positions enriched by the 7-plex OCEANS AML panel.

| Gene | Enrichment Region (GRCh38) |
|---|---|
| MAP2K1 | Chr15: 66,435,114-66,435,129 |
| MAP2K1 | Chr15: 66,436,814-66,436,830 |
| MAP2K1 | Chr15: 66,481,788-66,481,804 |
| MAP2K2 | Chr19: 4,117,540-4,117,558 |
| MAP2K2 | Chr19: 4,110,573-4,110,588 |
| MAP2K2 | Chr19: 4,101,089-4,101,106 |
| AKT1 | Chr14: 104,776,700-104,776,714 |
| AKT3 | Chr1: 243,695,699-243,695,726 |
| NRAS | Chr1: 114,716,123-114,716,137 |
| NRAS | Chr1: 114,713,894-114,713,912 |
| KRAS | Chr12: 25,245,346-25,245,358 |
| KRAS | Chr12: 25,227,328-25,227,346 |
| PIK3CA | Chr3: 179,218,291-179,218,309 |
| PIK3CA | Chr3: 179,234,293-179,234,307 |
| BRAF | Chr7: 140,753,333-140,753,353 |

TABLE S4-2. Chromosome positions enriched by the 15-plex OCEANS melanoma panel.

| Gene | Enrichment Region (GRCh38) |
|---|---|
| CTNNB1 | Chr3: 41,224,607-41,224,626 |
| CTNNB1 | Chr3: 41,224,645-41,224,663 |
| ARID1A | Chr1: 26,729,717-26,729,730 |
| AXIN1 | Chr16: 346,763-346,779 |
| TERT | Chr5: 1,295,114-1,295,123 |
| JAK1 | Chr1: 64,845,513-64,845,528 |
| PTEN | Chr10: 87,933,139-87,933,154 |
| TP53 | Chr17: 7,675,081-7,675,091 |
| TP53 | Chr17: 7,674,879-7,674,896 |
| TP53 | Chr17: 7,674,210-7,674,222 |
| TP53 | Chr17: 7,673,802-7,673,816 |

TABLE S4-3. Chromosome positions enriched by the 11-plex OCEANS HCC panel.

| Gene | Enrichment Region (GRCh38) |
|---|---|
| AKT1 | Chr14: 104,780,200-104,780,218 |
| ALK | Chr2: 29,222,334-29,222,352 |
| ALK | Chr2: 29,220,830-29,220,847 |
| ALK | Chr2: 29,213,992-29,214,009 |
| ALK | Chr2: 29,209,816-29,209,832 |
| BRAF | Chr7: 140,753,326-140,753,346 |
| BRAF | Chr7: 140,781,595-140,781,614 |
| DDR2 | Chr1: 162,778,599-162,778,613 |
| EGFR | Chr7: 55,174,001-55,174,015 |
| EGFR | Chr7: 55,174,769-55,174,790 |
| EGFR | Chr7: 55,181,309-55,181,322 |
| EGFR | Chr7: 55,181,378-55,181,391 |
| EGFR | Chr7: 55,191,817-55,191,831 |
| KRAS | Chr12: 25,227,341-25,227,356 |
| KRAS | Chr12: 25,245,340-25,245,352 |
| KRAS | Chr12: 25,225,609-25,225,628 |
| MAP2K1 | Chr15: 66,435,106-66,435,124 |
| MET | Chr7: 116,771,974-116,771,998 |
| NRAS | Chr1: 114,716,111-114,716,127 |
| NRAS | Chr1: 114,713,907-114,713,924 |
| PIK3CA | Chr3: 179,218,294-179,218,311 |
| PIK3CA | Chr3: 179,234,281-179,234,301 |
| PTEN | Chr10: 87,957,911-87,957,922 |
| ROS1 | Chr6: 117,318,223-117,318,238 |
| TP53 | Chr17: 7,675,081-7,675,091 |
| TP53 | Chr17: 7,674,879-7,674,896 |
| TP53 | Chr17: 7,674,217-7,674,230 |
| TP53 | Chr17: 7,673,802-7,673,816 |

TABLE S4-4. Chromosome positions enriched by the 28-plex OCEANS NSCLC panel.

| Gene | Mutation | Median EF |
|---|---|---|
| KIT | 2446G>C | 1420 |
| FLT3 | 2504A>T | 886 |
| DNMT3A | 2645G>A | 32.4 |
| IDH1 | 394C>T | 1280 |
| IDH2 | 418C>T | 128 |
| IDH2 | 515G>T | 61.7 |
| NPM1 | 863_864insTCTG | 361 |

TABLE S4-5. Median Enrichment Fold (EF) values observed experimentally for the 7-plex AML OCEANS panel.

| Gene | Mutation | Median EF |
|---|---|---|
| AKT1 | 235G>T | 5650 |
| AKT3 | 49G>A | 478 |
| BRAF | 1799T>A | 78.1 |
| KRAS | 34G>A | 625 |
| KRAS | 180_181GA>TT | 4220 |
| MAP2K1 | 169A>G | 534 |
| MAP2K1 | 361A>T | 221 |
| MAP2K1 | 607G>A | 202 |
| MAP2K2 | 169T>G | 811 |
| MAP2K2 | 373A>T | 192 |
| MAP2K2 | 619G>A | 2580 |
| NRAS | 34G>A | 143 |
| NRAS | 181G>T | 99.2 |
| PIK3CA | 1624G>A | 447 |
| PIK3CA | 3139G>A | 6570 |

TABLE S4-6. Median Enrichment Fold (EF) values observed experimentally for the 15-plex melanoma OCEANS panel.

#### Section S5. Mutations Detected in Melanoma Clinical Samples and NGS Comparison

**Comparison of NS and Illumina NGS data on clinical samples.** Fig. S5-1 shows comparison of OCEANS and Illumina NGS data separately for melanoma fresh frozen (FF) and FFPE clinical tissue samples. Mutations called by the OCEANS panel for melanoma, NSCLC and HCC clinical samples are listed in the excel file. Fig. S5-2 shows the location of TERT promoter mutation in a homopolymer region. FF and FFPE samples where insufficient amounts of DNA (< 5ng) were extracted were excluded from these lists, and OCEANS panels were not run on them. Table S5-1 shows clinical sample information for melanoma FF samples; FFPE sample clinical information were not available. Table S5-2 and Table S5-3 show clinical sample information for NSCLC and HCC FF and FFPE samples.

There are significantly higher number of mutations calls in FFPE samples compared to FF samples, such as the MAP2K1 c.371C>T mutation observed in 15 of the melanoma FFPE samples. The high concordance with digital PCR (Supplementary Section S6) generally suggests that these called variants are not due to systematic errors in the OCEANS method. A more likely explanation is that these mutations may have arisen from cytosine deamination DNA damage from FFPE treatment. This is supported by the fact that FFPE mutation calls had a significant over-representation of C>T and G>A mutations that derive from cytosine deamination.

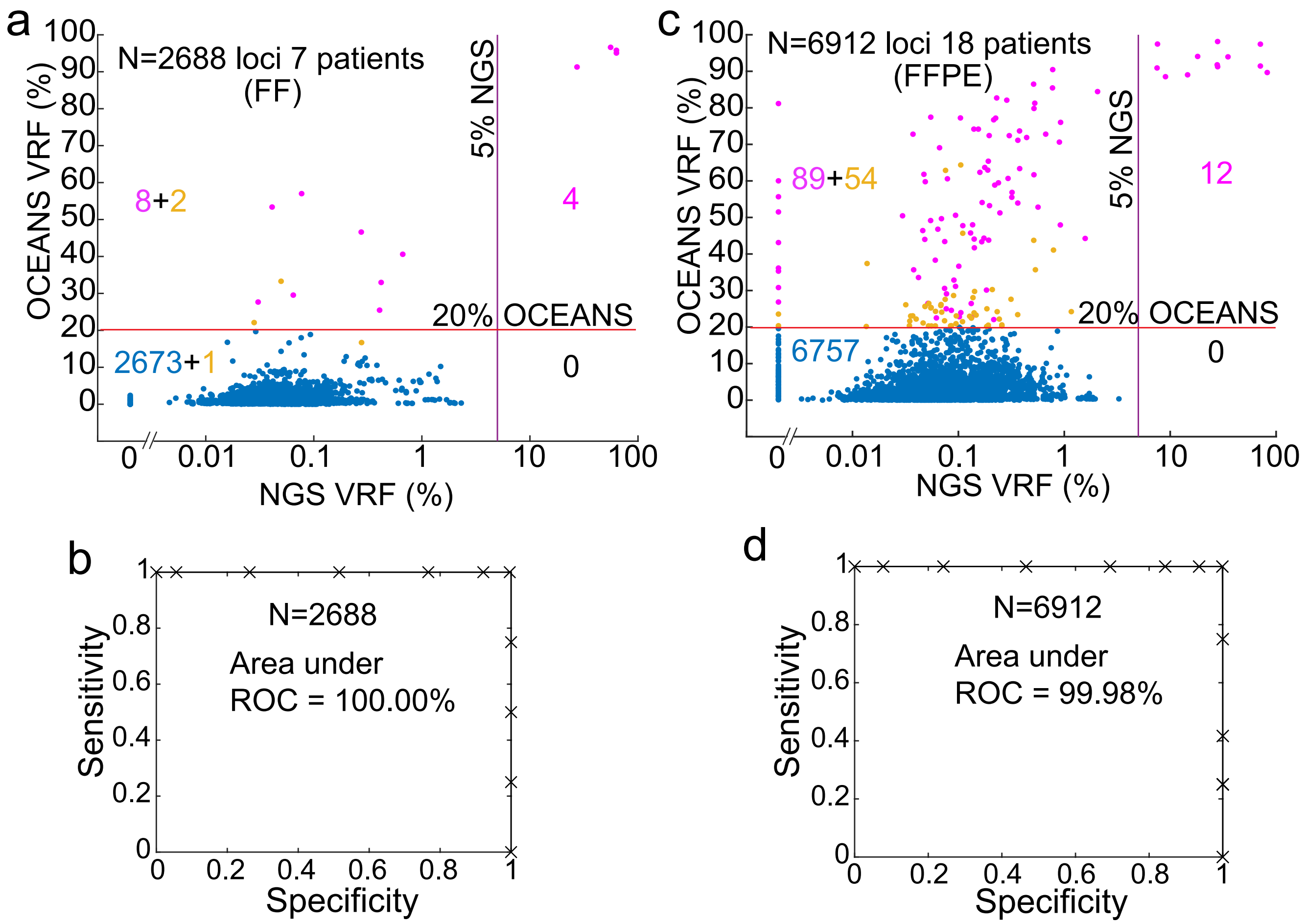


FIG. S5-1. Scatter plot comparison of NS and Illumina NGS data on melanoma clinical samples. **(a)** Comparison on fresh frozen clinical samples. %VRF for each nucleotide position in the enrichment region is plotted against the corresponding %VRF in NGS. Number of data points in each quadrant is indicated. 8 putative variants in the top-left quadrant had both VRF >20% and Clair score above 180 (purple dots). 3 putative variants had either VRF >20% or Clair score above 180 (yellow dots). **(b)** ROC curve for fresh frozen clinical samples. NGS inferred VAF > 5% were considered true positives. **(c)** Comparison on FFPE clinical samples. VRF for each nucleotide position in the enrichment region is plotted against the corresponding VRF in NGS. Number of data points in each quadrant is indicated. 89 putative variants in the top-left quadrant had both VRF >20% and Clair score above 180 (purple dots). 54 putative variants had either VRF >20% or Clair score above 180 (yellow dots). **(d)** ROC curve for FFPE clinical samples.

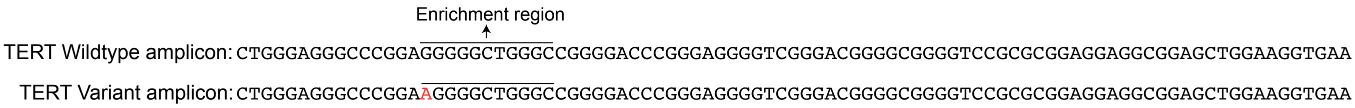


FIG. S5-2. TERT amplicon sequence in HCC OCEANS panel. G>A mutation is located within a homopolymer region in TERT promoter. Any variant in the underlined sequence is enriched by BDA.

| Sample | Stage | Age | Gender | Pathology |
|---|---|---|---|---|
| FF172 | IV | 60's | Female | Malignant melanoma |
| FF3 | IV | 40's | Female | Malignant melanoma |
| FF52 | IV | 60's | Male | Malignant melanoma |
| FF26 | IV | 50's | Male | Malignant melanoma |
| FF61 | IV | 40's | Female | Malignant melanoma |
| FF2 | IV | 40's | Female | Malignant melanoma |
| FF20 | II | 50's | Male | Malignant melanoma |

TABLE S5-1. Melanoma fresh/frozen clinical sample information.

| Sample | Stage | Age | Gender | Pathology |
|---|---|---|---|---|
| FFPE1 | IV | 70's | Male | Adenocarcinoma of lung |
| FFPE2 | IB | 40's | Female | Adenocarcinoma of lung |
| FFPE3 | IIB | 60's | Female | Adenocarcinoma of lung |
| FFPE4 | IB | 70's | Female | Carcinoma of lung, non-small cell |
| FFPE5 | IB | 70's | Male | Carcinoma of lung, squamous cell |
| FFPE6 | IIB | 70's | Male | Carcinoma of lung, squamous cell |
| FFPE7 | IB | 60's | Female | Adenocarcinoma of lung |
| FFPE8 | IIIA | 70's | Male | Carcinoma of lung, squamous cell |
| FFPE9 | IB | 80's | Male | Adenocarcinoma of lung, acinar, papillary |
| FFPE10 | IB | 70's | Male | Carcinoma of lung, squamous cell |
| FFPE11 | IB | 60's | Male | Carcinoma of lung, squamous cell |
| FFPE12 | IA | 60's | Female | Carcinoma of lung, squamous cell |
| FFPE13 | IIB | 60's | Female | Adenocarcinoma of lung |
| FFPE14 | IIIA | 60's | Male | Carcinoma of lung, squamous cell |
| FFPE15 | IB | 70's | Female | Carcinoma of lung, squamous cell |
| FFPE16 | IIB | 70's | Female | Adenocarcinoma of lung |
| FFPE17 | IB | 60's | Male | Adenocarcinoma of lung |
| FFPE18 | IIIA | 60's | Male | Adenocarcinoma of lung, papillary |
| FF19 | IA | 70's | Male | Adenocarcinoma of lung |
| FF20 | IA | 50's | Female | Adenocarcinoma of lung |
| FF21 | IA | 60's | Female | Carcinoma of lung, squamous cell |
| FF22 | IV | 50's | Male | Adenocarcinoma of lung |
| FF23 | IIIA | 70's | Male | Carcinoma of lung, squamous cell |

TABLE S5-2. NSCLC clinical sample information.

| Sample | Stage | Age | Gender | Pathology |
|---|---|---|---|---|
| FFPE1 | IIIB | 60's | Male | Hepatocellular carcinoma |
| FFPE2 | IB | 40's | Male | Hepatocellular carcinoma |
| FFPE3 | IB | 50's | Female | Hepatocellular carcinoma |
| FFPE4 | II | 70's | Male | Hepatocellular carcinoma |
| FFPE5 | IIIA | 40's | Male | Hepatocellular carcinoma |
| FFPE6 | IB | 60's | Male | Hepatocellular carcinoma |
| FFPE7 | IIIB | 40's | Female | Hepatocellular carcinoma |
| FFPE8 | IVA | 50's | Female | Hepatocellular carcinoma |
| FFPE9 | IB | 50's | Male | Hepatocellular carcinoma |
| FFPE10 | NA | 40's | Male | Hepatocellular carcinoma |
| FFPE11 | I | 50's | Male | Hepatocellular carcinoma |
| FFPE12 | NA | 40's | Male | Hepatocellular carcinoma |
| FFPE13 | NA | 60's | Male | Hepatocellular carcinoma |
| FFPE14 | NA | 60's | Male | Hepatocellular carcinoma |
| FFPE15 | IIIA | 60's | Male | Hepatocellular carcinoma |
| FFPE16 | I | 70's | Male | Hepatocellular carcinoma |
| FF17 | II | 60's | Male | Hepatocellular carcinoma |
| FF18 | IIIA | 80's | Male | Hepatocellular carcinoma |
| FF19 | IIIA | 80's | Male | Hepatocellular carcinoma |
| FF20 | IIIA | 70's | Male | Hepatocellular carcinoma |
| FF21 | II | 60's | Male | Hepatocellular carcinoma |

TABLE S5-3. HCC clinical sample information.

#### Section S6: ddPCR Comparison Experiments

We performed a total of 24 ddPCR comparison experiments on 6 FFPE DNA samples, testing 4 mutations each (BRAF p.V600E, KRAS p.G13D, KRAS p.E62K, and MAP2K1 p.P124L). We observe that 21 of the 24 matched experiments gave concordant results, with 10 of these concordant results being concordant positives. 2 of the 11 concordant negative values were deemed OCEANS negative because they only satisfied one of the two criteria (based on VRF and Clair), suggesting that the orthogonal bioinformatic validation improved variant call accuracy.

Individual ddPCR results and comparison to OCEANS % VRF are shown in figures S6-1, S6-2, S6-3 and S6-4. Green dots are HEX (WT) positive droplets, blue dots are FAM (Variant) positive droplets, red dots are double positive, and black dots are negative droplets. VAF was calculated as VAF = ((Variant positive+Double positive)/(WT positive + Variant positive+Double positive)).

| Sample | BRAF p. V600 | | KRAS p. G13D | | KRAS p. E62K | | MAP2K1 p. P124L | |
|---|---|---|---|---|---|---|---|---|
| | OCEANS | ddPCR VAF | OCEANS | ddPCR VAF | OCEANS | ddPCR VAF | OCEANS | ddPCR VAF |
| FFPE3 | Yes | 31.13% | Yes | 0.03% | No | 0% | Yes | 0.28% |
| FFPE13 | Yes | 0.66% | No* | 0% | No | 0% | Yes | 0.15% |
| FFPE17 | No | 0% | Yes | 0% | No | 0.43% | No | 0% |
| FFPE18 | No | 0% | Yes | 0.04% | No | 0% | No | 0.50% |
| FFPE19 | No | 0% | No* | 0% | Yes | 0.05% | Yes | 0.64% |
| FFPE25 | No | 0% | No | 0% | Yes | 0.02% | Yes | 0.14% |

TABLE S6-1. Summary of ddPCR results for BRAF p.V600E, BRAF p.V600K, KRAS p.G13D, KRAS p.E62K, MAP2K1 p.P124L mutations in melanoma clinical samples. Concordant positive results are displayed in green, concordant negative results in blue, and discordant results in red. *OCEANS variant call satisfied one condition for a variant call, either VRF > 20% or Clair score >180.

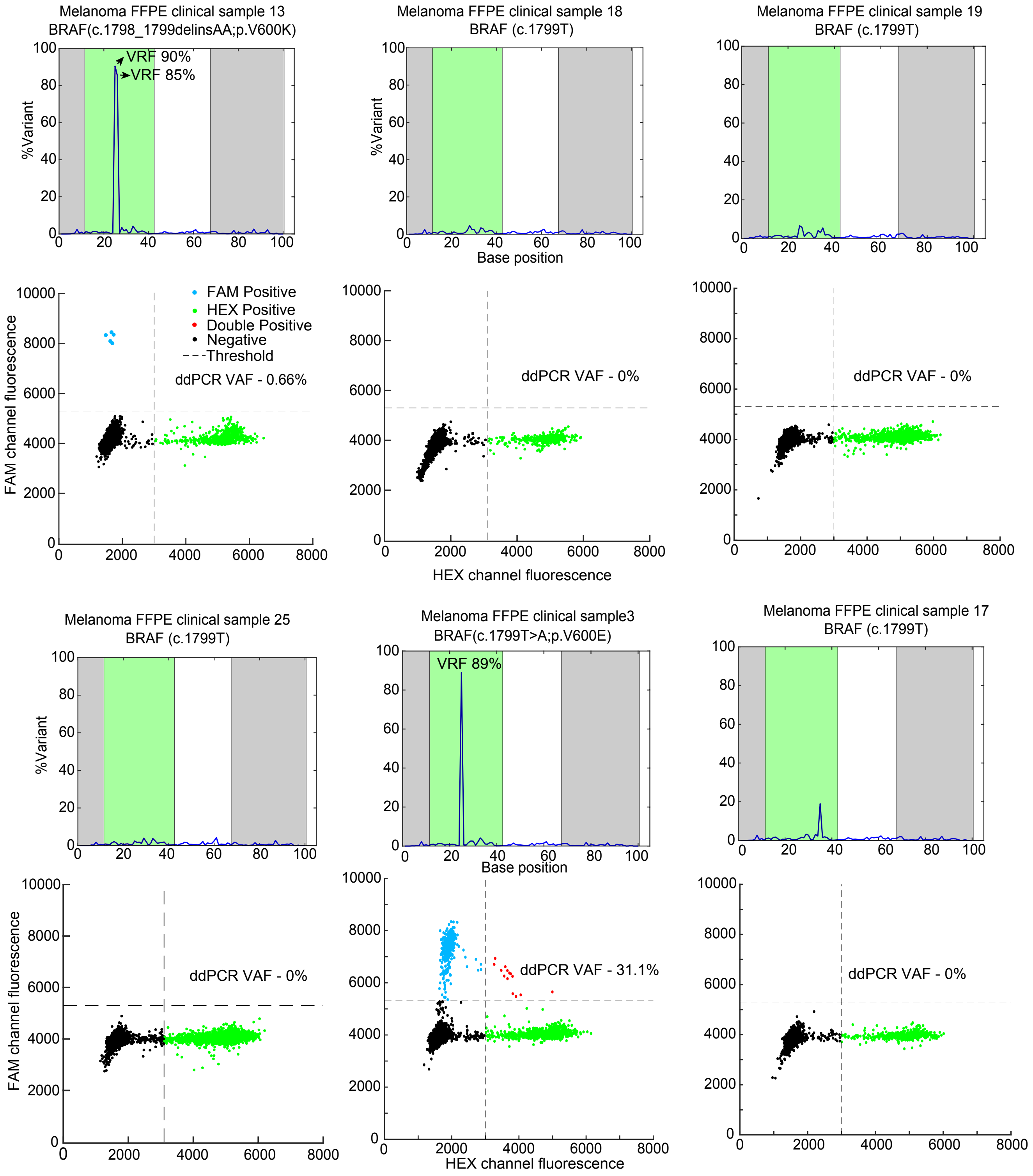


FIG. S6-1. NS and ddPCR comparison for mutation at BRAF (c.1799T) position in melanoma clinical samples. Top panel shows OCEANS results. Forward and reverse primer regions are shaded in gray, and the BDA enrichment region is shaded in green. Bottom panel shows ddPCR results for the same sample.

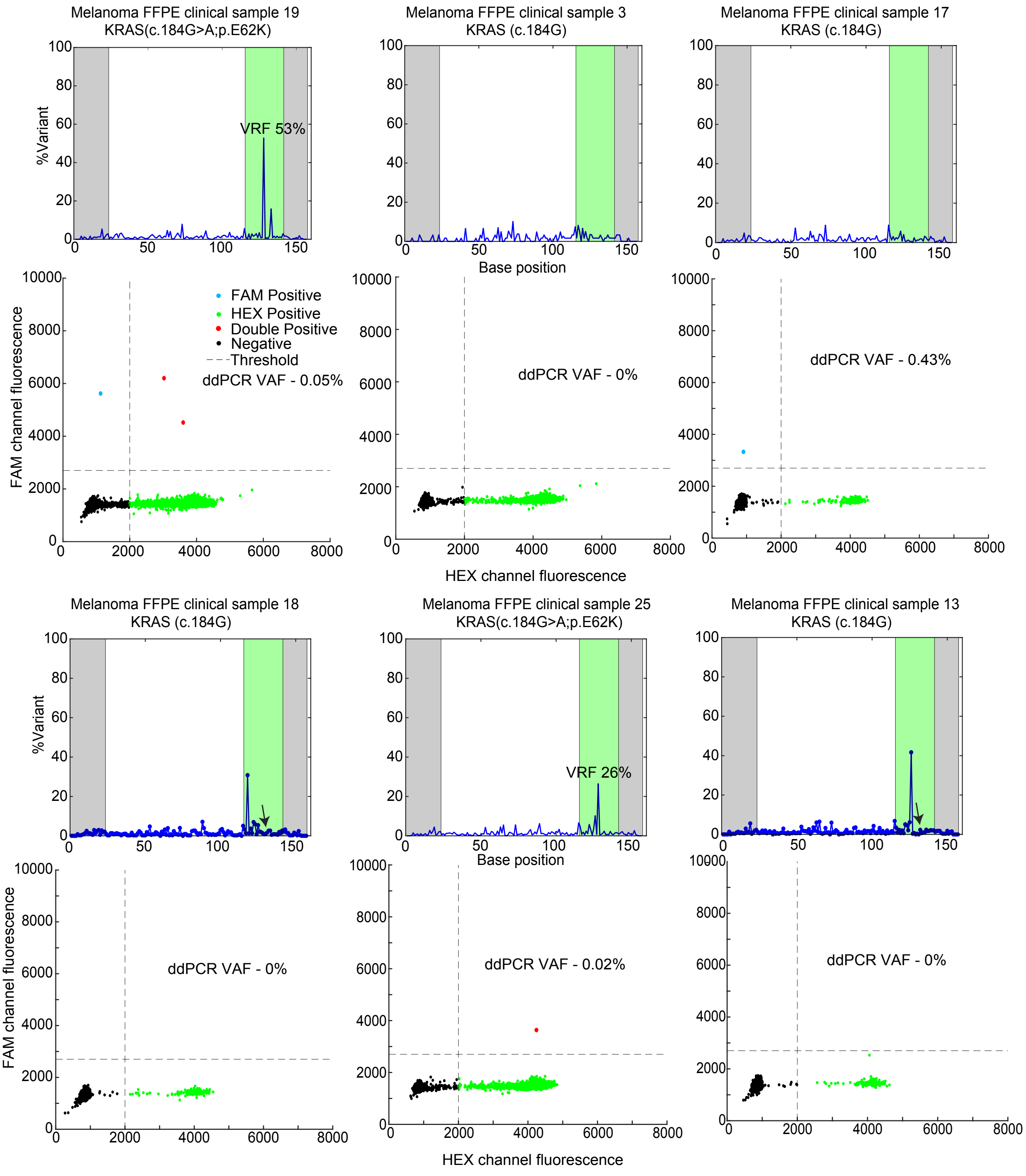


FIG. S6-2. NS and ddPCR comparison for mutation at KRAS (c.184G) position in melanoma clinical samples. Top panel shows OCEANS results. Forward and reverse primer regions are shaded in gray, and the BDA enrichment region is shaded in green. Bottom panel shows ddPCR results for the same sample. For FFPE sample 18 and 13, the c.184 position is indicated by arrows.

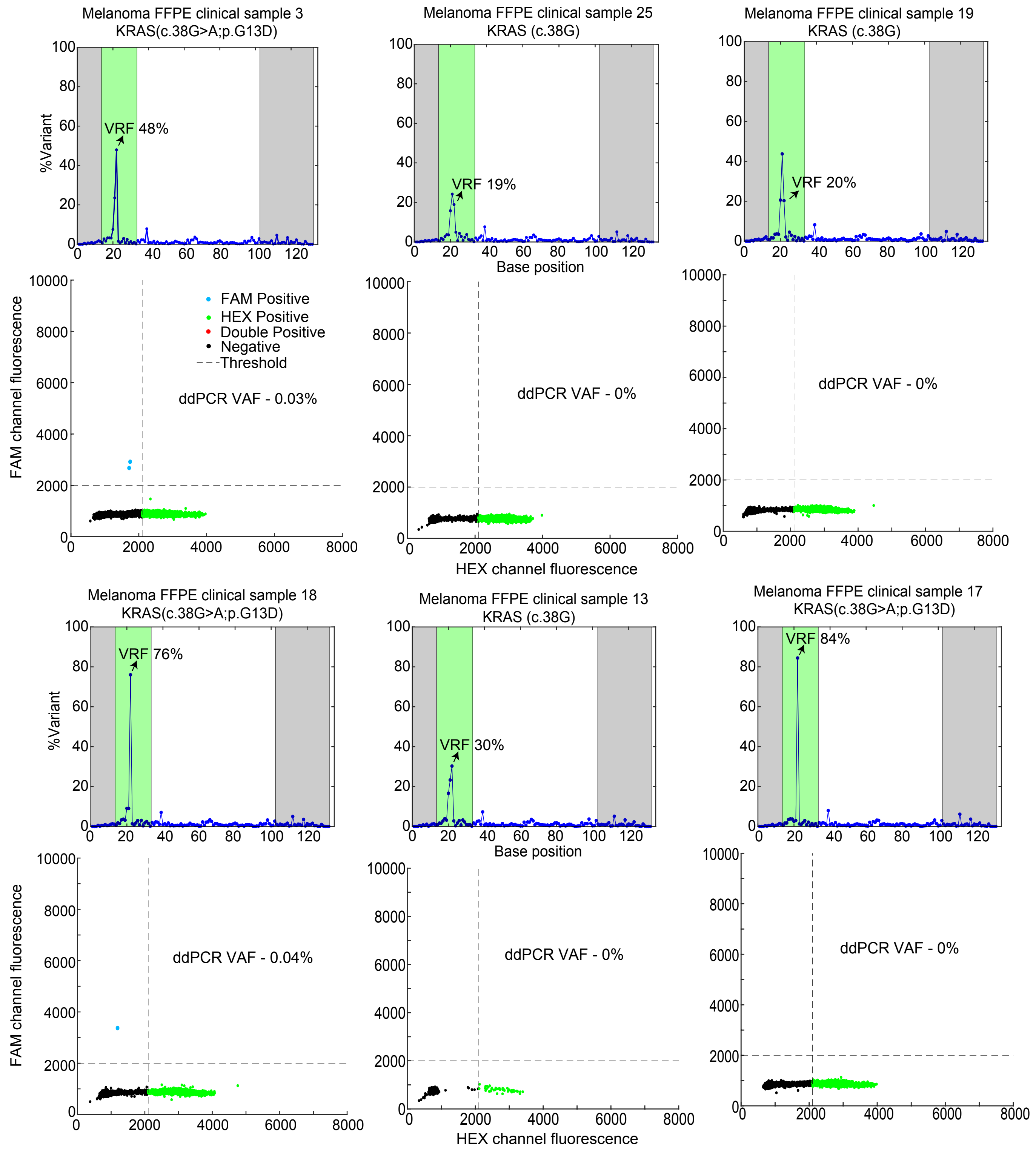


FIG. S6-3. NS and ddPCR comparison for mutation at KRAS (c.38G) position in melanoma clinical samples. Top panel shows OCEANS results. Forward and reverse primer regions are shaded in gray, and the BDA enrichment region is shaded in green. Bottom panel shows ddPCR results for the same sample. The c.38 position is indicated by arrows. The % VRF at this loci was high for all samples tested, but Clair scores were ¿180 only for FFPE samples 3, 18 and 17.

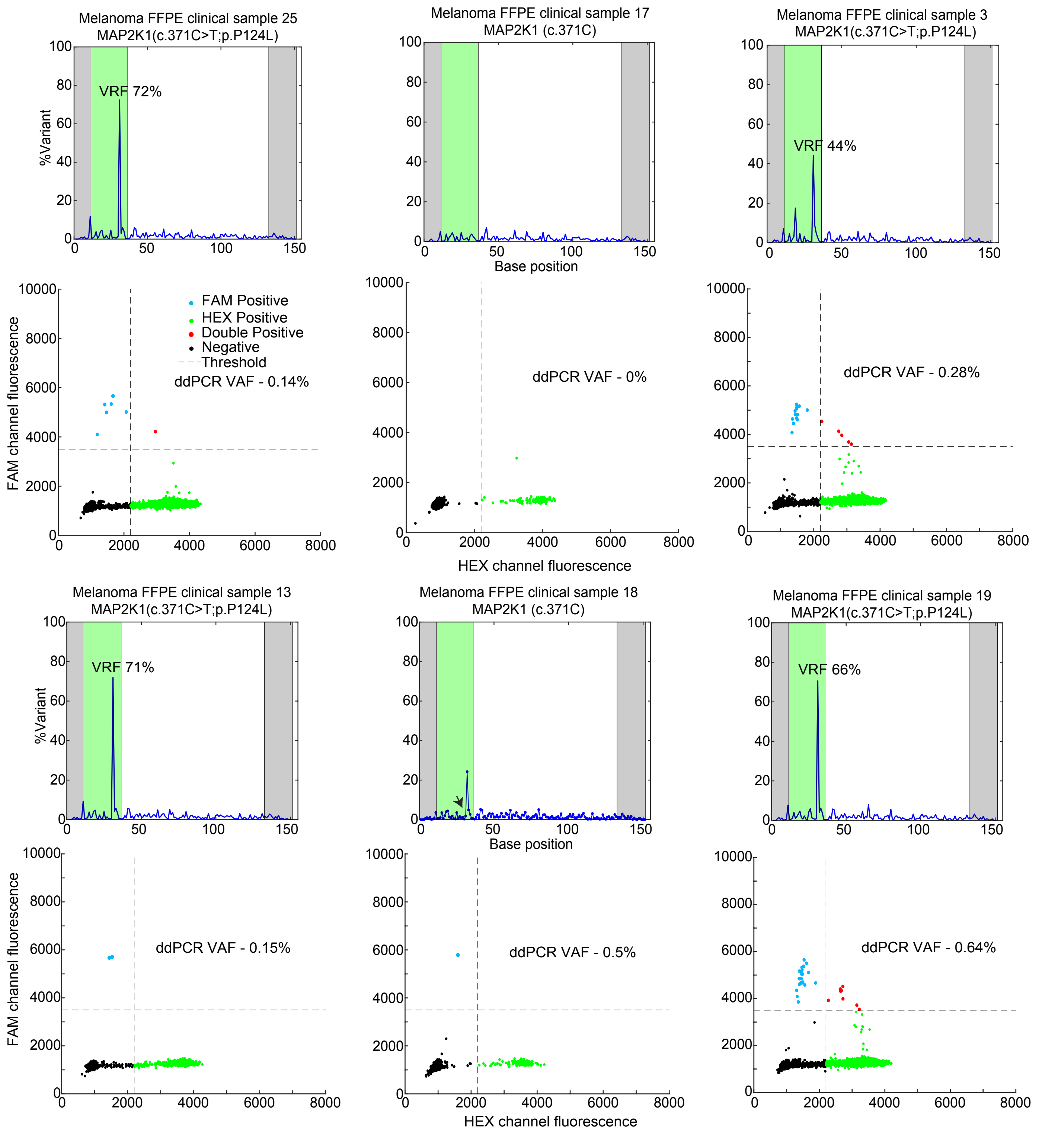


FIG. S6-4. NS and ddPCR comparison for mutation at MAP2K1 (c.371C) position in melanoma clinical samples. Top panel shows OCEANS results. Forward and reverse primer regions are shaded in gray, and the BDA enrichment region is shaded in green. Bottom panel shows ddPCR results for the same sample. The c.371 position is indicated by arrow for FFPE sample 18.
